# Deep Learning and Single Cell Phenotyping for Rapid Antimicrobial Susceptibility Testing

**DOI:** 10.1101/2022.12.08.22283219

**Authors:** Aleksander Zagajewski, Piers Turner, Conor Feehily, Hafez El Sayyed, Monique Andersson, Lucinda Barrett, Sarah Oakley, Mathew Stracy, Derrick Crook, Christoffer Nellåker, Nicole Stoesser, Achillefs N. Kapanidis

**Affiliations:** Department of Physics, University of Oxford, Parks Road, Oxford, OX1 3PJ, United Kingdom; Kavli Institute for Nanoscience Discovery, University of Oxford, South Parks Road, Oxford OX1 3QU, United Kingdom; Nuffield Department of Medicine, University of Oxford, John Radcliffe Hospital, Oxford, OX3 9DU, United Kingdom; Department of Microbiology and Infectious Diseases, Oxford University Hospitals NHS Foundation Trust, Oxford, OX3 9DU, United Kingdom; Sir William Dunn School of Pathology, University of Oxford, South Parks Road, Oxford, OX1 3RE, United Kingdom; Nuffield Department of Women’s & Reproductive Health, University of Oxford, Big Data Institute, Oxford, OX3 7LF, United Kingdom

## Abstract

The rise of antimicrobial resistance (AMR) is one of the greatest public health challenges, already causing up to 1.2 million deaths annually and rising. Current gold-standard antimicrobial susceptibility tests (ASTs) are low-throughput and can take up to 48 hours, with implications for patient care. We present advances towards a novel, rapid AST, based on the deep-learning of single-cell specific phenotypes directly associated with antimicrobial susceptibility in *Escherichia coli*. Our models can reliably (80% single-cell accuracy) classify untreated and treated susceptible cells, across a range of antibiotics and phenotypes - including phenotypes not visually distinct to a trained, human observer. Applying models trained on lab-reference susceptible strains to clinical isolates of *E. coli* treated with ciprofloxacin, we demonstrate our models reveal significant (p<0.001) differences between resistant and susceptible populations, around a fixed treatment level. Conversely, deploying on cells treated with a range of ciprofloxacin concentrations, we show single-cell phenotyping has the potential to provide equivalent information to a 24-hour growth AST assay, but in as little as 30 minutes.

## INTRODUCTION

Antimicrobial resistance (AMR) is a major public health challenge, causing an estimated 1.2 million deaths annually ^1^, with this number predicted to rise much further if left unchecked. AMR represents the evolutionary effect of antimicrobial selection pressures in the context of the short bacterial cell cycle, leading to adaptation by natural selection through a variety of molecular mechanisms ^2^. Several clinical strategies to address the AMR crisis have been considered. One strategy relies on the continuous development of novel antimicrobial agents to outpace bacterial evolution; however, this strategy alone is neither scientifically nor economically viable ^3,4^. Another strategy relies on the conservation of the existing antimicrobial arsenal through strict stewardship and regulation, which remains challenging to implement universally ^5^. A third option, as part of stewardship, is through diagnostic improvements, including more rapid Antimicrobial Susceptibility Testing (AST) methods, which allow better tailoring of antibiotic treatment regimens given to patients ^6^. A combination of these and other approaches, such as vaccination, is likely needed, as no single strategy currently represents a complete solution across all settings.

Existing ASTs provide phenotypic quantification of the Minimum Inhibitory Concentration (MIC) of an antibiotic of choice for isolates cultured from infected patients, and can be complemented by targeted nucleic-acid assays for known resistance determinants. AST may also be preceded by species identification (e.g., using MALDI-ToF). The major drawback of current ASTs is their relatively slow speed and throughput, requiring culture-based isolation of clinical pathogens, expert operators and laboratory space, and ∼24 hours turnaround time from sampling ^7^. Initial antimicrobial regimens given to sick patients are therefore usually broad-spectrum, which may maximise collateral patient-level effects such as perturbation of gut flora, and contribute to the selection and dissemination of AMR at both the patient- and population-levels.

Multiple novel approaches to improve the speed of AST exist, including biosensors, genomic assays, and hybridization approaches ^8^; however, most of these remain in the development stage ^9^, and have not yet been translated into practice. Many of these methods assess an *entire* bacterial culture, and the lack of single-cell specificity leaves them insensitive to heterogeneity in cell populations, such as the presence of persister cells. A potential solution to this problem is the use of single-cell specific ASTs, which address the heterogeneity problem while also offering higher throughput by evaluating the effect of antimicrobials *directly* on cells, rather than relying on secondary markers, such as growth of an entire culture. Multiple such candidate ASTs have been proposed, enabled by platforms such as flow cytometry ^10^, Raman spectroscopy ^11,12^, fluorescent probes in droplet microfluidic devices ^13^, impedence cytometry ^14^ and others. Further opportunity in single-cell ASTs comes from their integration with widefield microscopy ^15–18^, which further increases throughput by enabling real-time simultaneous monitoring of large numbers of individual cells. Such cellular imaging produces rich, high-volume, unstructured data that are well suited to machine-learning based analysis, and in particular, by modern deep-learning techniques. These techniques have been used to great effect to produce AST inference models from genomic ^19,20^ and metabolomic ^21–24^ data, where their ability to execute their own feature engineering maximises the usage of complex, unstructured data. With similar insights, deep-learning has been applied to widefield microscopy to produce candidate ASTs that provide phenotypic quantification by monitoring single-cell growth ^25,26^ or motion patterns ^27,28^ in the presence of antibiotics.

Another microscopy approach relies on directly evaluating the effect of antimicrobials on cellular structures, such as the bacterial nucleoid or the cell membrane. These structures have been characterized experimentally and computationally ^29,30^, and were used as single-cell phenotypes to profile cytological pathways to understand the mode of action of antibiotics ^31^, and to test for methicillin resistance in *Staphylococcus aureus* ^32^. A wide range of nucleoid and cell membrane cytological phenotypes under different treatment conditions have since been established for a range of Gram-positive and Gram-negative species ^33–35^. Such a phenotyping approach has notable advantages over single-cell microscopy assays monitoring growth or motion patterns: results are available on the timescale of a single bacterial life cycle rather than several lifecycles, the method is applicable to difficult-to-culture pathogens, and there is no requirement for continuous tracking of individual live cells over time. However, cellular phenotyping may be affected by phenotypic plasticity, whereby small genotypic and environmental differences can strongly influence the displayed phenotype.

In this work, we introduce a novel, single-cell, microscopy-based AST that addresses many important shortcomings of existing assays. We combine single-cell phenotypes of the nucleoid and cell membrane with modern Convolutional Neural Networks (CNNs) to design an AST based on deep phenotyping of individual cells that display different physiological responses to antibiotics. CNNs feature a hierarchical pattern of learnable convolution filters, which allows efficient learning in heterogenous imaging data without manual feature engineering – removing the main bottleneck of previous work ^32^. Our Deep Antimicrobial Susceptibility Phenotyping (DASP) platform uses widefield micrographs to rapidly classify antibiotic-treated cells as either susceptible or resistant. We have developed specific models for four antibiotics, each representative of an antibiotic family with a different mode of action: the fluoroquinolone ciprofloxacin (which targets DNA synthesis), the aminoglycoside gentamicin (targets protein synthesis), the beta-lactam co-amoxiclav (targets cell-well synthesis) and rifampicin (targets RNA synthesis). We show our models are robust to phenotypic plasticity by training them on a lab strain of *Escherichia coli* and then deploying them successfully on several *E. coli* clinical isolates with different MICs, where we were able to distinguish reliably susceptible and resistant isolates around a fixed treatment concentration. Furthermore, we demonstrated that varying the treatment concentration generates a dose-response relationship that might allow precise quantification of the MIC of isolates an order of magnitude faster than established gold-standard clinical techniques.

## RESULTS

### Detecting antibiotic susceptibility based on deep learning of single-cell subcellular phenotypes

We designed an AST that takes as an input bacterial cultures grown in rich medium until a consistent optical density and then treated with an antibiotic of choice for a time sufficient to produce a phenotypic change (e.g., in the nucleoid morphology) to the given antibiotic in the case the strain is susceptible. Upon cell fixation and staining (Fig. 1A), antibiotic-resistant cells that remain unaffected by the treatment should display a resistant phenotype, which highly resembles the original untreated phenotype (see Fig. S1 for examples comparing the resistant and untreated phenotypes in resistant clinical isolates). In contrast, antibiotic-susceptible cells will show visually distinct phenotypic changes – a distinct, antibiotic-specific, susceptible phenotype (Fig. 1A, left).

**Figure 1.**
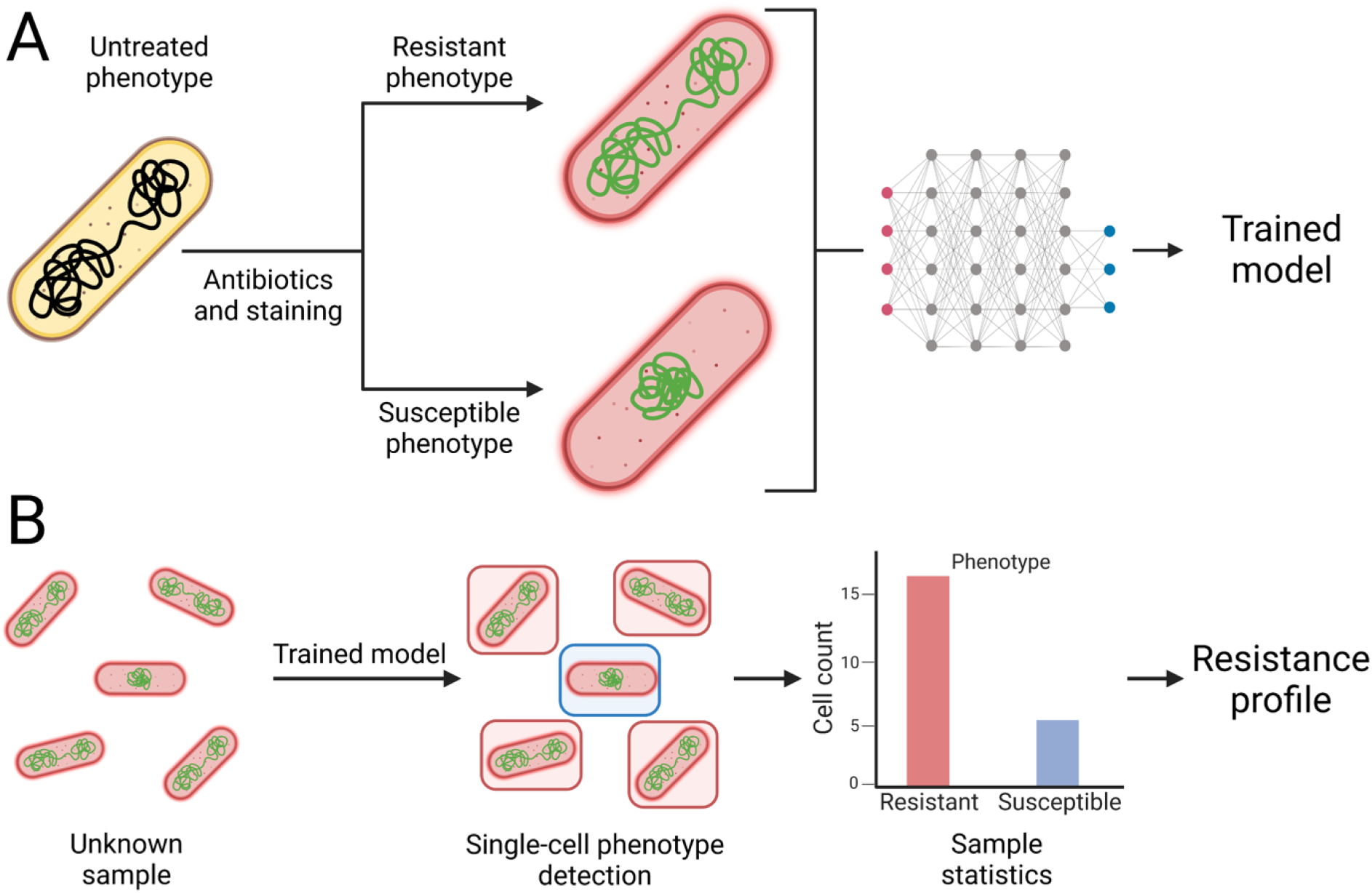
AST assay based on single-cell phenotyping and deep-learning. **A**. Live *E. coli* cells are treated with an antibiotic, inducing changes in subcellular morphology. Susceptible cells show strong phenotypic changes associated with the effect of antibiotic action, creating a distinct susceptible phenotype. Resistant cells are not affected by the antibiotic – the resistant phenotype is similar to the untreated phenotype. Cells are fixed, and nucleoids and cell membranes are fluorescently stained. The sample is imaged under a widefield fluorescence microscope. A deep learning pipeline is trained to distinguish the susceptible phenotype from the resistant (untreated) phenotype, with single cell resolution. **B**. An unknown sample can be processed and fed into the trained model, which classifies the phenotypes on a single cell level to produce sample-wide classification statistics. These statistics can then be used to obtain information on the resistance of the entire sample.

After collecting a large number of micrographs (each representing an image containing 50-200 cells), individual cells representing either the untreated or susceptible phenotype were used to train segmentation and classification models. At test time, the trained models segment micrographs of treated cells and classify individual cells into one of the two categories with regards to antibiotic susceptibility; examining the distribution of classifications reports on the resistance of the entire unknown sample (Fig. 1B).

### Generating antibiotic-resistant and antibiotic-susceptible phenotypes

To implement the concept above, we characterized the untreated phenotype of lab reference *E. coli* strain MG1655, as well as the susceptible phenotypes of the same strain to four clinically relevant antibiotics: ciprofloxacin (CIP), gentamycin (GENT), rifampicin (RIF) or co-amoxiclav (COAMOX) (see *Methods* and Fig. S2). To capture the untreated phenotype (and use it as a proxy for the resistant phenotypes, where we expect no treatment-induced changes in the nucleoid and membrane) and the susceptible phenotype for each antibiotic, we stained the bacterial nucleoid with the DNA-binding fluorophore DAPI (green signals in Fig. 2A-E), and the membrane with the lipid stain Nile Red (NR; red signals in Fig. 2A-E), revealing the organization of the nucleoid and overall cell morphology as a function of treatment.

**Figure 2.**
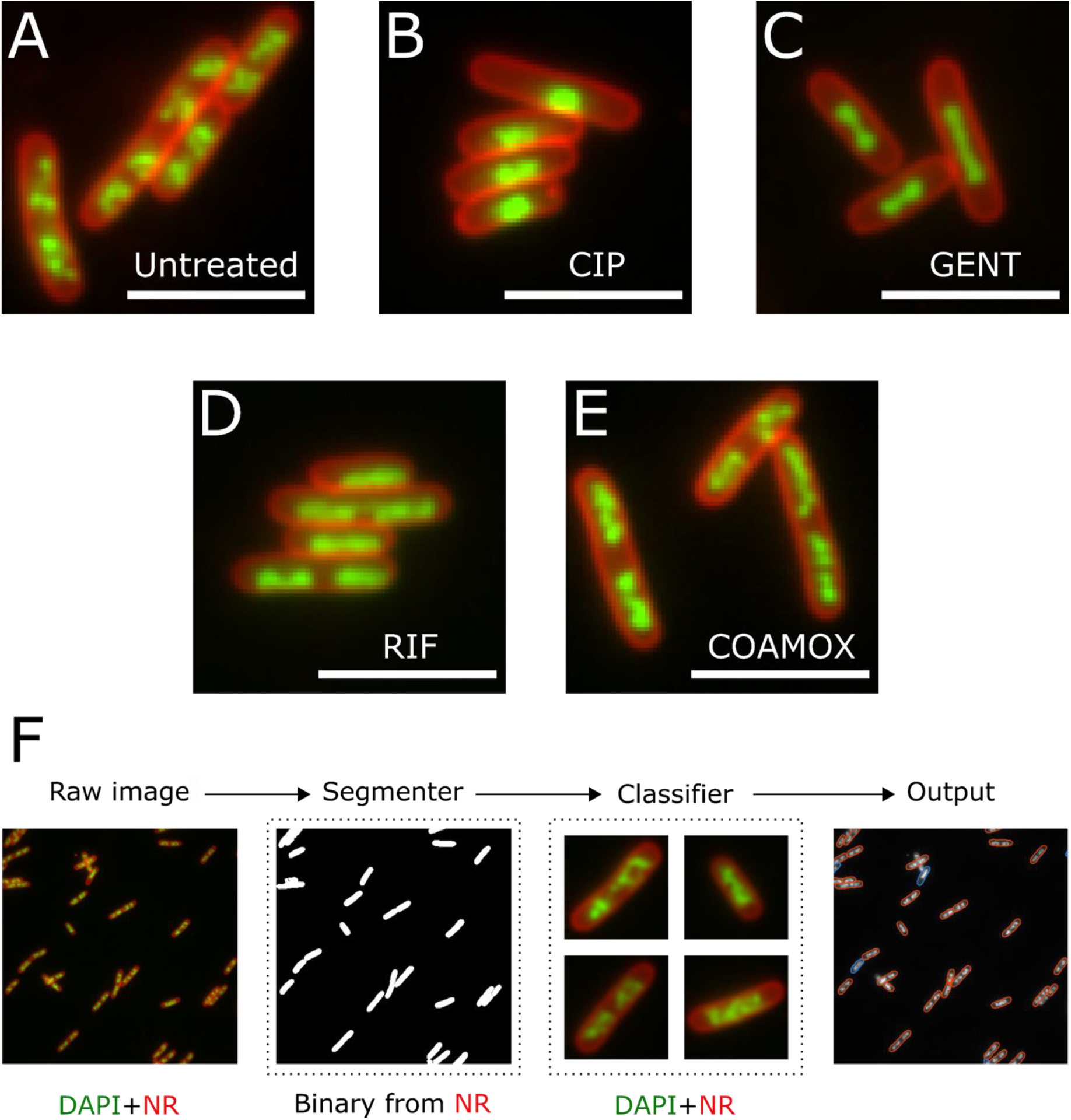
Segmentation and classification pipeline. **A**. Untreated phenotype in the MG1655 *E. coli* strain, which resembles the resistant phenotype. Green, nucleoid stained with DAPI; red, membrane stained with Nile Red. Scale bar, 5 μm. **B**. As in A, but for the ciprofloxacin susceptible phenotype. **C**. As in A, but for the Rifampicin susceptible phenotype. **D**. As in A, but for the Gentamicin susceptible phenotype. **E**. As in A, but for the Co-amoxiclav susceptible phenotype. Note that this phenotype has more subtle differences from the untreated phenotype. **F**. A multichannel image consisting of DAPI (green) and Nile Red (red) is split into individual channels. A Mask-RCNN segmenter segments single-instance binary masks from the Nile Red channel. Binary masks are used to isolate single cells. A phenotype classifier classifies individual cells into either the resistant or susceptible phenotype, using both channels.

In the untreated phenotype (Fig. 2A), distinct copies of the chromosome were seen in each cell, organized into heterogenous macrodomains by nucleoid-associated proteins ^36^. In contrast, incubating susceptible MG1655 with ciprofloxacin produced a compaction of the chromosome towards the cell center due to topoisomerase IV inhibition ^37^ (Fig. 2B). Similarly, incubation with gentamicin, which binds to the 30S ribosomal subunit and interferes with translation elongation, also leads to nucleoid compaction, although the chromosomes in this case do not merge fully into one spot (Fig. 2C). Incubation with rifampicin, which inhibits transcription initiation by RNA polymerase, led to decompaction of the nucleoid (Fig. 2D); individual chromosomes could still be distinguished, but the macrodomains were lost. Finally, exposure to co-amoxiclav produces a subtle susceptible phenotype – whilst some differences in the organization of the macrodomains can be seen, the effect is more challenging to discriminate visually from the untreated phenotype.

Representative full fields of view for all phenotypes are provided in Figs. S3 and S4. To classify the phenotypes, we designed a 2-stage deep-learning pipeline (Fig. 2F). In the first stage, a Mask-Region based Convolutional Neural Network (RCNN) model ^38^ segments individual cells from whole micrographs using the image generated using NR stain. In the second stage, a separate DenseNet121 classifier classifies cells into phenotypes using the images generated using both NR and DAPI stains. Together, both stages allow precise quantification of the sample response to antibiotic treatment with single-cell resolution.

### Single-instance cell segmentation by Mask-RCNN

To segment cells from micrographs, a Mask-RCNN model was built on a ResNet50 backbone, and trained for a 2-class segmentation task (cell/background) on an ensemble dataset of annotated micrographs of treated and untreated cells from 6 repeat imaging experiments of MG1655 (Fig. S5C), consisting in total of 29,297 ground truth, manually curated cells in 459 fields of view. During training, the model was continually validated on a validation set, consisting of 9,044 cells in 115 fields of view.

We evaluated the performance of our segmentation on a dataset consisting of all the micrographs across all treatments in a “holdout” experiment – from which no cells or micrographs were used either in training or hyperparameter optimization. Evaluating a total of 155 micrographs containing 13,247 ground-truth, manually curated cells, we detected a total of 12,147 cells (92% maximum total recall). To quantify the quality of the segmentation, we calculated the average precision-recall curves at a range of Intersection over Union (IoU) thresholds, as well as the associated segmentation confidence (Fig. S6). The precision-recall curve quantifies the tradeoff between precision (i.e., the fraction of returned results that are relevant), and recall (i.e., the fraction of total relevant results that were successfully returned), whereas the IoU quantifies the area overlap between the detection instances and ground truth instances needed to count as a successful detection – as the IoU threshold increases, the task becomes harder. We achieved an Average Precision (AP) of 85.5% at the standard IoU threshold of 0.5; the AP decreased slightly to 79.3% at the stricter IoU threshold of 0.75.

### Distinguishing resistant and susceptible single-cell phenotypes

To classify segmented cells into distinct phenotypes, we trained DenseNet121 ^39^ classifiers in a range of computational experiments (Fig. S5B-C), for the binary classification of resistant and susceptible single-cell phenotypes generated using MG1655 and one classifier per antibiotic. As discussed above, treated MG1655 cells were used to generate the susceptible class, whilst untreated cells were used to generate the resistant class.

First, we trained each classifier on an ensemble dataset, where all untreated and treated cells from 6 experiments were combined into one dataset from which cells for the training, validation and test sets were drawn randomly without replacement (see *Methods*); this “ensemble” approach provides the expected upper bound for model performance. Second, to examine the performance loss from the upper bound due to experimental variation within the training data, we performed an experimental cross-validation: one of the 6 experiments was withheld for testing, and the model trained on a class-balanced dataset consisting of an equal number of cells drawn randomly from each of the remaining experiments. This “cross-validation” experiment was rotated around with the final reported result being the sum over 6 different models trained and tested on different permutations of experiments. Lastly, to evaluate the robustness of the classifiers against experimental variation, we trained a model on a class-balanced dataset of equal numbers of cells drawn randomly from the 6 experiments, and then evaluated it on a class-balanced dataset of the same equal number of randomly selected cells from a 7^th^, “holdout” experiment. Notably, no data from the holdout experiment was used for training, or hyperparameter optimization of any of the classifiers.

With this procedure, we achieved an excellent (>84%) single-cell classification accuracy in the holdout experiments across all antibiotic conditions (Fig. 3), with comparable statistics in the other computational experiments; full numerical results across all experiments and antibiotics are provided in Fig. S7. Untreated cells, which were used to generate the resistant class, were predominantly classified as resistant (i.e., not displaying a physiological response to an antibiotic), whilst treated cells were classified as susceptible (i.e., displaying the expected physiological response).

**Figure 3.**
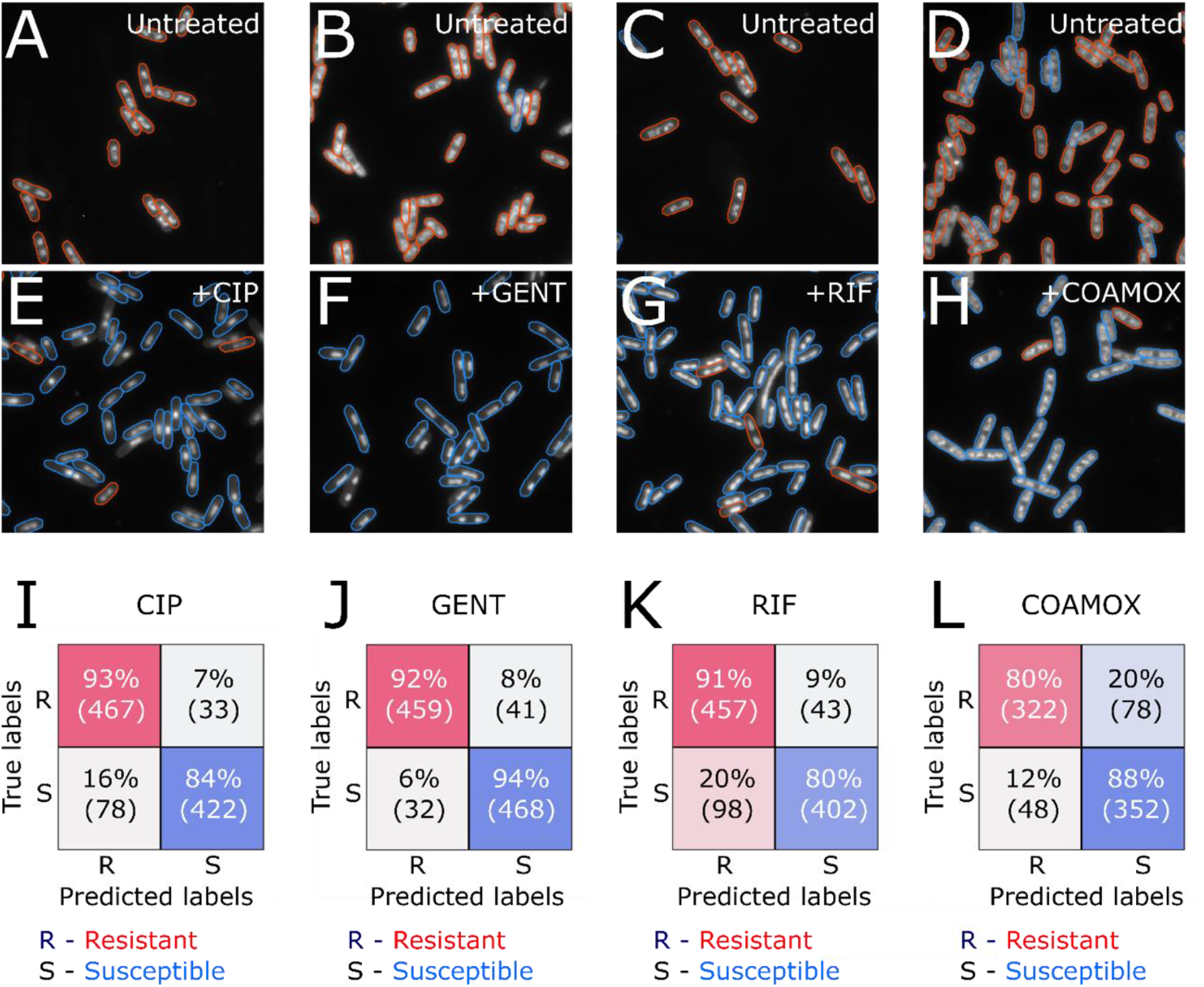
Binary classification of resistant and susceptible phenotypes in *E. coli* MG1655 strain. **A**. Representative field of view (FoV) of untreated cells shown in grayscale, with an overlay showing phenotype detections. Resistant classifications shown in red, susceptible ones shown in blue. Evaluation carried out with the ciprofloxacin resistant/susceptible classifier. **B-D**. As in A, but with the gentamycin classifier (panel B), the rifampicin classifier (panel C), or the co-amoxiclav classifier (panel D). **E**. As in A, but with ciprofloxacin-treated cells. **F**. As in B, but with gentamicin-treated cells. **G**. As in C, but with rifampicin-treated cells. **H**. As in D, but with co-amoxiclav-treated cells. **I**. Holdout test performance of the ciprofloxacin classifier, evaluated on class-balanced, randomly sampled 1000 cells from an independent holdout experiment, from which no data was drawn for model training or hyperparameter optimization. Percentage and absolute class counts shown in both the resistant (R) and susceptible (S) classes. **J**. As in I, but for the gentamicin classifier. **K**. As in I, but for the rifampicin classifier. **L**. As in I, but for the co-amoxiclav classifier, and with a dataset of 800 cells.

To understand the decision-making process of the CNN classifier, and the potential failure modes, we used saliency mapping ^40^ to produce attention heatmaps over example single-cell phenotype inputs; such mapping highlights the pixels that contribute most to the classification decision. We observed that, for all antibiotics, in correctly classified cells, the classifier focuses primarily on nucleoid structure and organization, with some attention given to the membrane, as expected (Fig. S8). The same pattern was observed in cells that were misclassified (Fig. S9) – such cells did not show the full expected phenotype due to cell-to-cell heterogeneity. For example, some ciprofloxacin susceptible cells did not show a full nucleoid compaction during the treatment window, and were thus classified as resistant.

### Single-cell classification informs of resistance relative to treatment concentration in clinical isolates

Having validated both our segmentation and classification models on the MG1655 *E. coli* strain used to generate the training data, we deployed the models on six clinical isolates of *E. coli* (EC1-6), each with a different degree of resistance to ciprofloxacin (as exemplified by the MIC of each isolate, which we measured; see *Methods*) and linked to a resistance genotype derived from sequencing (Fig. S10). To evaluate the response of the isolates to ciprofloxacin, we used our segmentation model (trained on the ensemble dataset), and the binary antibiotic susceptibility classifier (resistant vs susceptible to ciprofloxacin) used to analyse holdout samples as described in the previous section.

Samples of clinical isolates were prepared using two conditions: applying no antibiotic treatment (untreated), or treating cells with ciprofloxacin at the same concentration and duration as the susceptible MG1655 used for training (20x EUCAST ^41^ breakpoint concentration [i.e., 10 mg/L], 30 min incubation). These samples underwent the same processing as the training samples, producing collections of micrographs for each sample. Those collections of micrographs were segmented to identify individual cells, and then the classification model was used to classify these cells as either resistant, or susceptible.

Prior to any antibiotic treatment, cells of all clinical isolates were predominantly classified correctly (i.e., in the resistant class – untreated cells showing no response to the antibiotic) and with high confidence; for example, we obtained 77% and 96% resistant classifications for untreated EC2 and EC5 (Fig. 4A, left, and Fig 4B, left, respectively). On the other hand, treated cells in clinical isolates susceptible to ciprofloxacin at the level used (that is, with an MIC below the 10 mg/L treatment concentration), displayed the susceptible phenotype, and were strongly classified as such, with 91% of all cells being classified as susceptible in the ciprofloxacin-susceptible EC2 (Fig. 4A, right).

**Figure 4.**
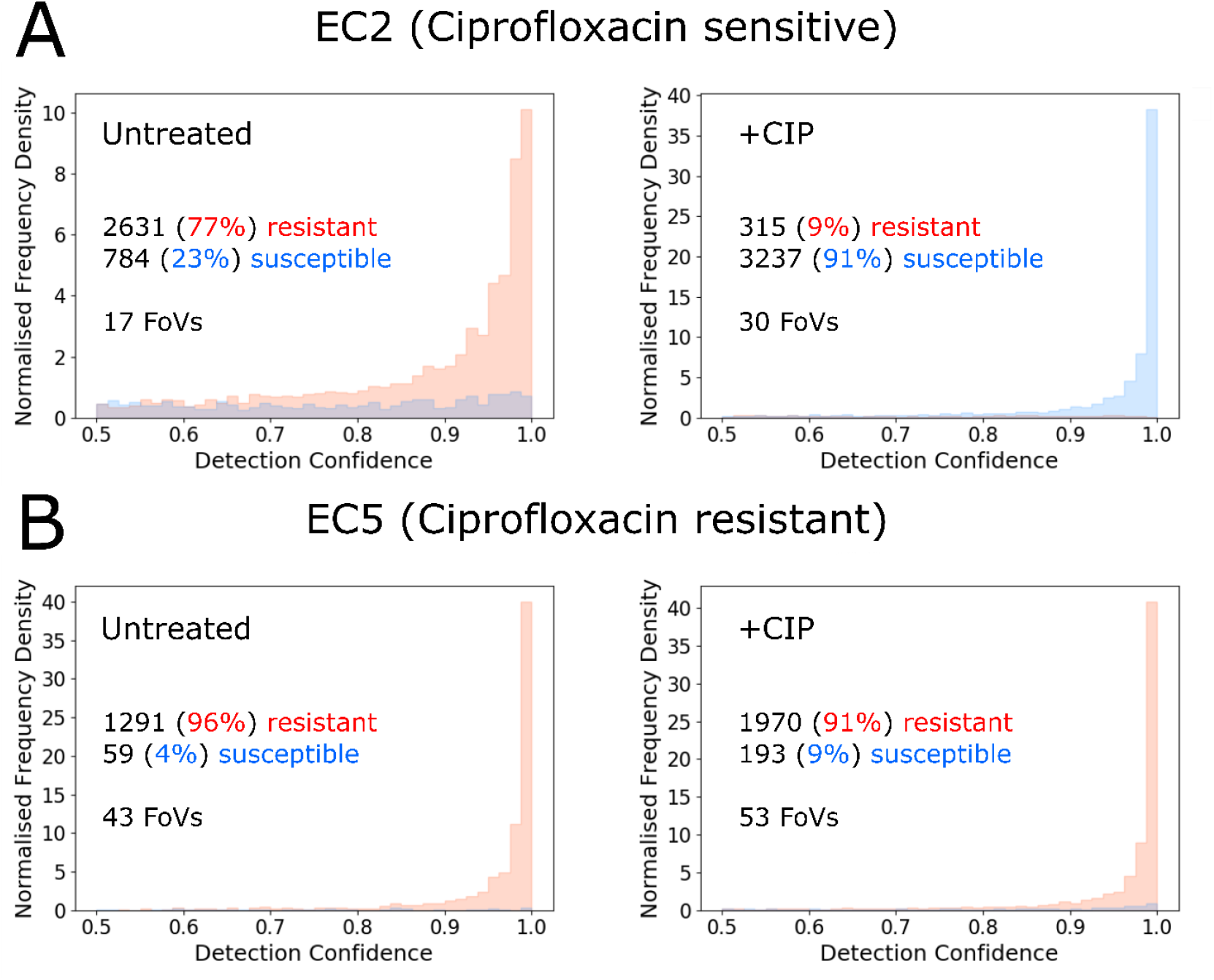
Distribution of single-cell classifications is sensitive to the resistance of clinical isolates. A. <u>Left</u>: histogram of detections from a representative imaging experiment of untreated, susceptible *E. coli* isolate EC2 cells, normalised to the total sum of detections, as a function of classification confidence. Percentages and absolute cell counts in each class indicated, together with the number of fields of view in the experiment (FoV). <u>Right</u>: as before, but for ciprofloxacin treated EC2 cells. **B**. As in A, but for ciprofloxacin-resistant EC5.

In contrast, for isolates with MICs above the treatment concentration, the large majority of treated cells were classified as resistant. For example, 91% of all cells were classified as resistant in treated but ciprofloxacin-resistant EC5, as expected (Fig. 4B, right).

This pattern was maintained across the library of all six clinical isolates (Fig. 5; see also Fig. S11-13 for representative fields of view and overlays showing phenotype detections, and Fig. S14 for the total number of cell detections in each repeat of each isolate and treatment condition). In isolates with MICs below the training and treatment concentration (EC1-4), there was a statistically significant (p-value < 0.001) increase in the ratio of susceptible detections in treated samples, as compared to the untreated samples. In sharp contrast, for resistant isolates with MICs above the training and treatment concentration (EC5-6), there was no statistically significant difference in the ratio of susceptible detections between treated and untreated samples. The size of the ratio of susceptible cells appeared correlated with the difference between the MIC and treatment concentration. Highly susceptible EC1-3 were characterized by showing 85% of susceptible cells in treated samples, whilst EC4 (MIC = 8mg/L, treatment concentration = 10mg/L) showed that only 38% of treated cells were classified as susceptible.

**Figure 5.**
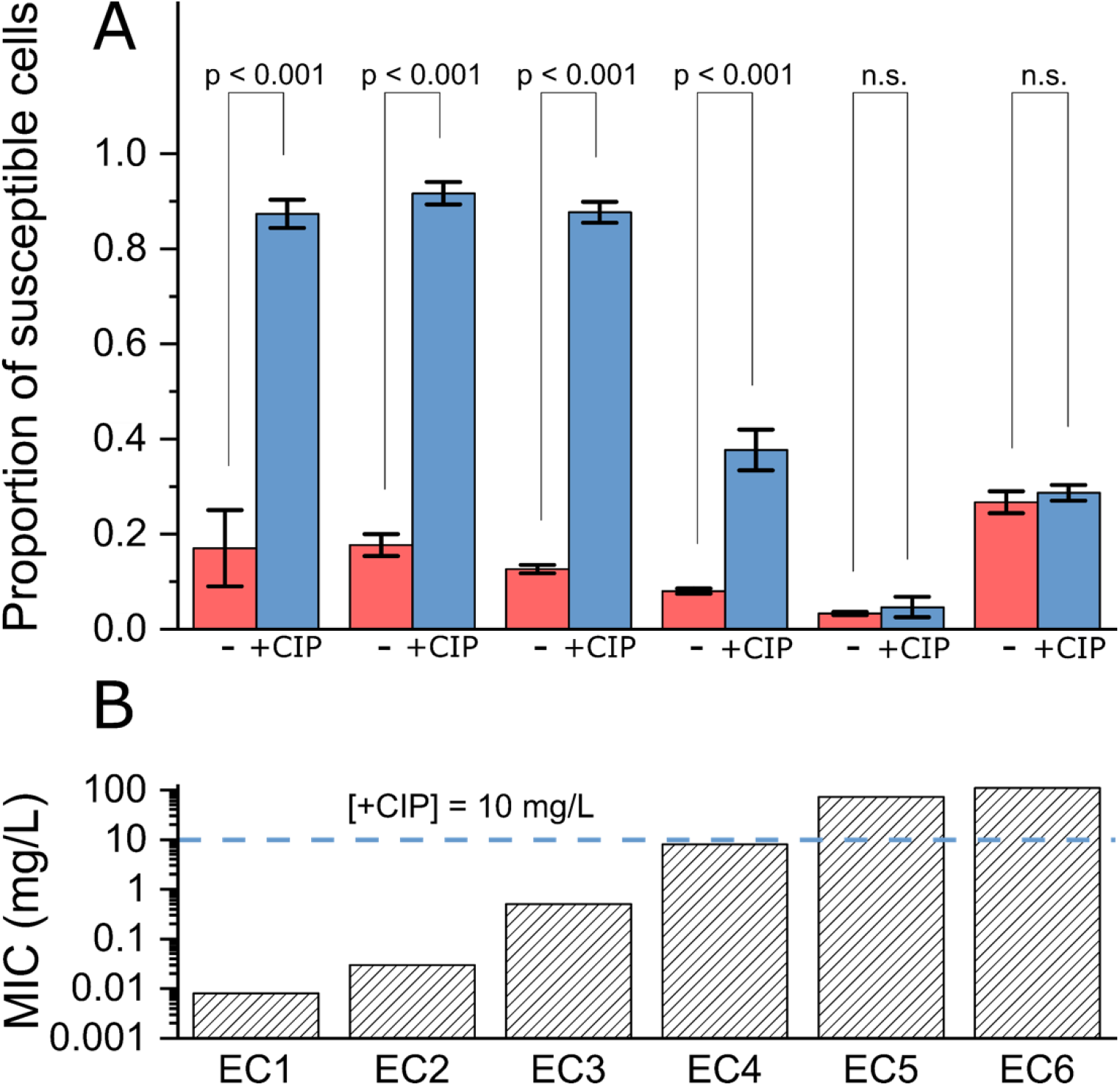
Changes in the overall fraction of susceptible cells upon treatment correlate with the degree of resistance in clinical isolates. **A**. Ratio of susceptible phenotype detections across different clinical isolates, in untreated (-) and ciprofloxacin treated (+CIP) samples. Average of 3 biological replicates; error bars show the standard error of the mean. Overlay shows Tukey range test p-values, carried out pairwise between corresponding untreated and treated sets of repeats (n=3), test carried out at significance level of 0.05, “n.s.” indicates not significant. **B**. Ciprofloxacin MICs of the clinical isolates, derived experimentally as described in *Methods*. Horizontal line indicates the treatment concentration used in the treated samples, which was used for both clinical isolates and MG1655 training data.

### Single-cell phenotyping provides AST information equivalent to a 24-hour growth assay, in 30 minutes

After demonstrating that our models can differentiate between clinical isolates resistant and susceptible to a fixed concentration of ciprofloxacin, we investigated the ciprofloxacin concentration dependence of our classifications, since we reasoned that it may report on the MIC value for different isolates. This time, we generated samples of three of the clinical isolates with different MICs (EC1, EC3 and EC5), treated them at different ciprofloxacin concentrations for 30 min, imaged them, and phenotyped them (see Fig. S15 for numbers of cell detections in each biological replicate of each isolate and treatment concentration).

Across all clinical isolates, cells treated at sub-MIC concentrations did not show a significant shift away from the resistant phenotype, and the ratio of cells classified as susceptible was low. Conversely, at concentrations in excess of the MIC, a strong response was observed, with >90% of the cells converting to the susceptible phenotype. At intermediate concentrations, the magnitude of the response varied logistically between the asymptotes; to quantify this relationship, we fitted asymmetric dose-response models to data (Fig. 6; see also *Methods*).

**Figure 6.**
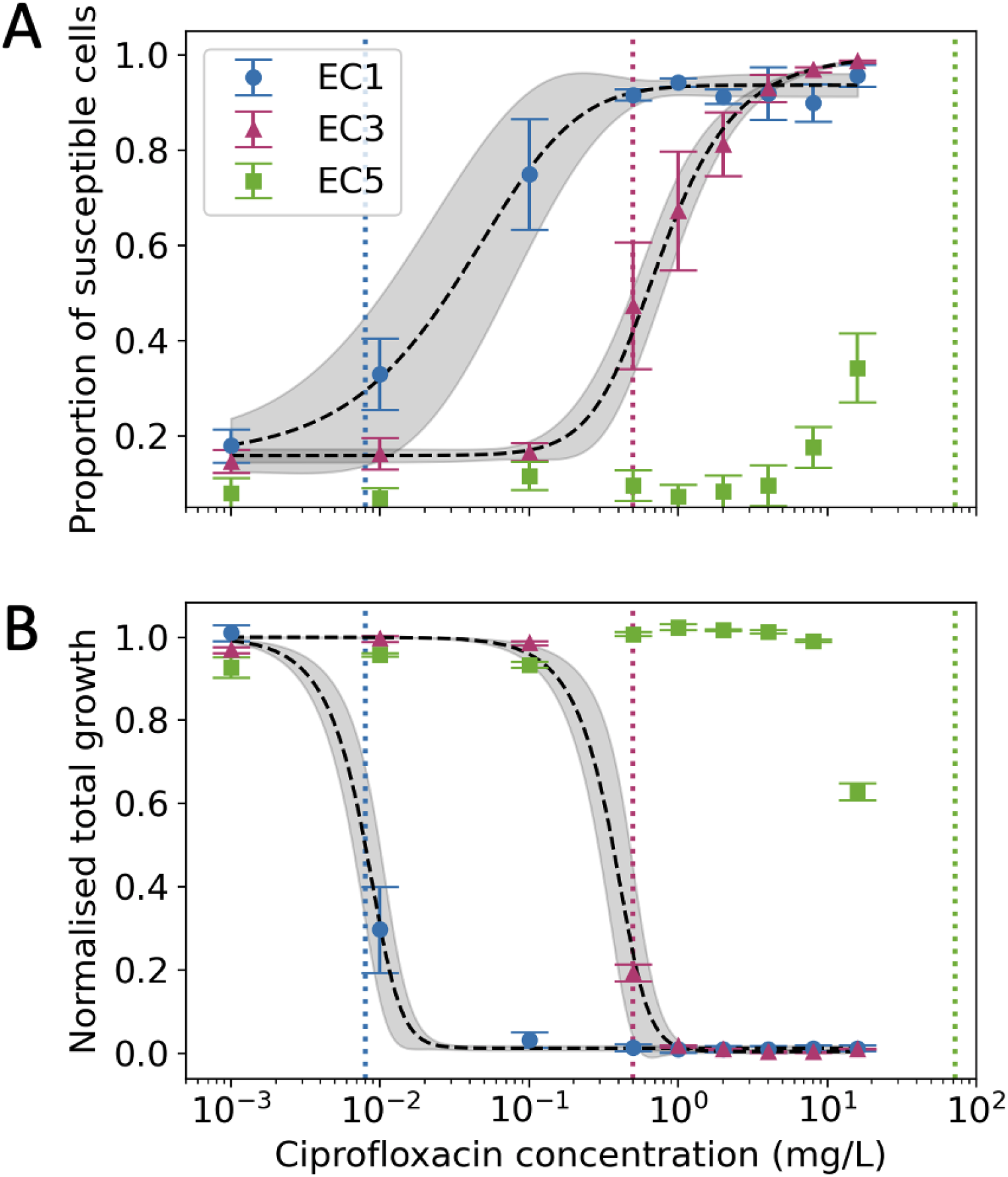
Single-cell phenotyping provides equivalent information to a 24-hour growth assay, but within 30 minutes. **A**. Ratio of susceptible detections as a function of ciprofloxacin treatment concentration. Average of three biological replicates, error bars show the standard error of the mean. Vertical coloured lines show the ciprofloxacin MICs of the isolates used. Black lines show the dose-response fit, grey regions show the 95% confidence band of the fit. **B**. Total bacterial growth in liquid culture over 24 hours in the presence of ciprofloxacin, normalised to the growth of untreated cells. Measured MICs of the isolates are: EC1 – 0.008 mg/L, EC3 – 0.5 mg/L, and EC5 – 72 mg/L.

Our phenotyping method provided results after as little as 30 min of treatment, whilst the gold-standard clinical AST relies on growth (or lack thereof) in the presence of antibiotics over much longer periods of time. To compare our result against the gold-standard, we measured growth curves of the isolates in the presence of ciprofloxacin over 24 hrs. From these curves, we calculated the total cell growth by numerical integration of the time-resolved optical density (OD_600_) signal, and normalized it to the growth of untreated cells, thereby creating a ratio of total cell growth as a function of ciprofloxacin concentration, relative to untreated cells (Fig. 6B). Across all isolates and ciprofloxacin concentrations, the total cell growth followed a relationship reciprocal to that of the ratio of susceptible detections. At sub-MIC concentrations, cell growth was not inhibited, leading to a high cell growth ratio, whilst at concentrations in excess of the MIC, no growth occurred. Again, at intermediate concentrations, a logistic, dose-response relationship was observed which mirrored the observed ratio of susceptible phenotypes.

In both dose-response models, the inflection points of the curves (corresponding to free parameter *c* in the model fit) were close to the MIC values of the isolates when measured using a routine diagnostic AST assay (see *Methods*). Specifically, in EC1, the measured MIC was 0.008 mg/L, with a corresponding inflection point at 0.011±0.001 mg/L; further, in EC2, the measured MIC was 0.5 mg/L, with curve inflection at 0.49±0.18 mg/L. Our results clearly establish that our approach can provide valuable MIC-related information for different clinical isolates, faster than established methods, whilst eliminating the time-consuming cell-culture step in the phenotypic susceptibility assay.

## DISCUSSION

By combining fluorescence imaging and deep-learning, we demonstrate our DASP platform provides robust, rapid single-cell antimicrobial susceptibility phenotyping for *E. coli* across a range of antibiotics with different mechanisms of action. DASP is fully compatible with analysing clinical isolates with different susceptibility to clinically relevant antibiotics (as we have shown for ciprofloxacin), and can provide equivalent information to a traditional growth assay in a small fraction of the time, while offering richer clinical information than current, colony-level, assays.

### A robust deep-learning phenotypic platform for advanced antibiotic susceptibility analysis

The high accuracy statistics across our experiments establish that our method is robust against both experimental variability and phenotype plasticity exhibited by different clinical isolates, showing substantial promise for the development of a robust clinical diagnostic assay. Considering that distinct nucleoid and membrane phenotypes have been identified a wide range of treatments and organisms ^33–35^; our method of susceptibility phenotyping should be applicable to a wide variety of antimicrobials and bacterial species.

Deploying our models on clinical isolates shows that the models can distinguish between resistant and susceptible isolates around a fixed treatment point, which can facilitate their use for decision-making around standard clinical breakpoints used for resistance classification, such as EUCAST breakpoints. Our comparison with the gold-standard growth assays also established that our approach can provide equivalent information regarding the MIC of a certain isolate in as little as 30 minutes (instead of several hours), limited only by the physiological response rate of the bacteria themselves, and is therefore well suited to the goal of a rapid assay.

Our assay may also offer a route towards a more detailed clinical definition of the MIC value, which is currently only defined by growth. Notably, the MIC value is emerging as an important independent factor in clinical management; e.g., co-amoxiclav MICs greater than 32 mg/L have recently been specifically associated with poor outcomes in *E. coli* bloodstream infections ^42^. Better and faster definition of bacterial MICs may therefore be relevant to optimizing treatment strategies and outcomes for individual patients.

### Comparison with other assays

Compared to current assays, our technique can also serve as a richer source of clinical information. Currently established ASTs operating on the colony level only offer aggregate, sample-wide information measured through secondary markers correlated with resistance, such as culture growth or genomic information. Compared to other candidate single-cell ASTs, DASP offers the potential for even fast timescales by allowing simultaneous interrogation of large numbers of single-cells (in contrast to previous cytometry or Raman approaches, which only interrogate one cell at a time), and does not require cell tracking (in contrast to previous widefield microscopy approaches focusing on single-cell growth or motion).

Previous single-cell phenotypic studies have implemented linear transformations and manual analysis of engineered features to classify methicillin susceptible and resistant *S. aureus* cells ^32^. Our approach represents a step-change over the approach above, since it transforms the assessment of antibiotic susceptibility into a non-linear classification of resistant phenotypes against susceptible phenotypes, addressed by CNNs. Using learnt features as opposed to engineered ones is advantageous, since it generalizes the technique by allowing subtle phenotypic changes to be detected (as seen in our co-amoxiclav results), and reducing human bias. In contrast to a more manual analysis, CNNs offer automatic processing of large volumes of data, scaling better with the aim of a rapid, robust assay.

### Future extensions

Our approach can be further extended to become more scalable and offer a faster time-to-result in the clinic. Currently, the assay operates on cultured clinical isolates, and thus does not yet remove the time-consuming laboratory steps needed to isolate and grow microorganisms from patient specimens. The isolates are cultured to a constant OD_600_ of ∼0.2 prior to processing, translating to ∼10^8^ Colony Forming Unit (CFU) counts - a count much higher than the CFU counts encountered in infected physiological body fluids ^43^. We envision that use of microfluidics will be instrumental in bypassing the pathogen isolation and culture steps by isolating and concentrating bacteria from patient specimens.

Our deep-learning approach relies on explicit classification of phenotypes; whilst that removes the need for engineered features, enables high-throughput and reduces human bias, it still requires that phenotype classes are homogenous across cells and isolates. Whilst true in our work, further work is required to validate this approach with more species and antibiotics. Our current approach also requires specific models to be trained for each combination of antibiotic and species, which may not scale well with the size of the problem space; however, a reformulation of the computational task should produce solutions that scales better. Finally, our assay can be coupled with species identification which can be performed using various methods, such as targeted FISH staining ^26^.

## METHODS

### Bacterial strains and sample preparation

The reference laboratory strain was *E. coli* MG1655. Clinical isolates were blood culture isolates of *E. coli* processed for diagnostic purposes and stored by the Microbiology Laboratory of the Oxford University Hospitals NHS Foundation Trust, Oxford, UK. Individual colonies of *E. coli* were cultured overnight in 5 ml of Lysogeny Broth (LB) at 37°C, then diluted 1:100 by volume in 5 ml of EZ Rich Defined Medium (RDM; Teknova) and cultured at 37°C until reaching an OD_600_ of ∼0.2 in a shaking incubator. Subsequently, 1 ml aliquots of the culture were treated with one of the antibiotics (Fig. S2) at the concentration and duration listed to produce cells showing the antibiotic-specific susceptible phenotype; aliquots of the culture were treated similarly but in the absence of an antibiotic in order to produce the resistant phenotype. Cells were then fixed by incubation in 2.5% formaldehyde solution for 30 min. Cells were washed 3 times with PBS via centrifugation at 4,500 RCF for 3 min, and then incubated with 100% ethanol for 10 min. Cells were resuspended in PBS and stained by adding 10 μg/L DAPI as the nucleic acid stain (GeneTex, catalogue number GTX16206) and 1 μg/L Nile Red as the membrane stain (Fisher Scientific, catalogue number 10464311) and incubated at room temperature for 10 min. Cells were washed twice with PBS and suspended in a small volume of 10-20 μl.

For ciprofloxacin titration assays, the antibiotic was serially diluted across a concentration range from 16 mg/L to 0.001 mg/L and co-incubated with 1 ml aliquots of bacteria as described above. Stained and prepared samples were imaged by mounting on agarose pads. Agarose pads were prepared consisting of 1% high-purity agarose (Bio-Rad, catalogue number #1613101) in half-concentration PBS solution, and imaged inverted through a glass slide that had been burned in a plasma cleaner at 500°C for 60 min.

All clinical isolates in the study had been whole-genome-sequenced on the Illumina platform as described previously ^44^, and AMR genotypes were assigned using the ResFinder ^45^ database with Abricate v0.9.8 ^46^ (--min-id 95 --min-cov 95). The MICs of the clinical isolates were calculated empirically by E-test strip, or where the MIC exceeded the range of the strip, by a broth dilution.

### Bacterial growth curves

Individual colonies of each strain were grown overnight at 37°C in LB broth and subsequently diluted to OD_600_ of ∼0.04 (1:100 dilution) in RDM. These cells were added in equal volume to a microtiter plate containing a prepared 2x dilution range of ciprofloxacin in RDM to a final volume of 200 μl. Inoculated plates were incubated at 37°C in a Tecan Sunrise plate reader, with an OD_600_ reading recorded at 15-min intervals, following a 5-sec orbital shaking. The same measurement was taken for a blank sample, consisting solely of the growth medium. To calculate total cell growth (Fig. 6B), the time-resolved OD_600_ signals were integrated numerically in time. From this, the integrated blank signals were subtracted, and finally all measurements were normalized to the growth of untreated cells by dividing each measurement by the measurement coming from untreated cells.

### Imaging

Agarose-mounted samples were imaged on a Nanoimager-S fluorescence microscope (Oxford Nanoimaging). Briefly, a blue (405 nm) and a green (532 nm) laser were combined using a dichroic mirror and coupled into a fibre optic cable. The fibre output was focused into the back focal plane of the objective (100x oil immersion, NA 1.4) Fluorescence emission was collected by the objective, separated into two emission channels and imaged onto a sCMOS camera (Orca flash V4, Hamamatsu). To make best use of the camera dynamic range DAPI signal was imaged using 405 nm excitation and Nile Red signal was imaged using 532 nm excitation; both signals were acquired consecutively. To ensure reproducibility, laser powers were kept constant at 1.5 kW/m^2^. For each of the two channels, for each field of view (FoV), a stack of 30 frames was acquired at 30 ms exposure and 33 Hz frequency. To automate the task and reduce human bias, the multiple acquisition capability of the microscope was used, and the microscope autofocused on each FoV prior to acquisition.

### Deep Learning: segmentation

To generate training data for the Mask-RCNN ^38^ segmenter, only the Nile Red channel of every FoV acquired was used. The 30 frames of each FoV were averaged to generate a grayscale image, which was further expanded to RGB space by replicating the grayscale image in each colour channel. The raw images were augmented on-the-fly by random cropping to a size of 256 by 256 pixels, followed by a random sequence of transformations including horizontal and vertical flips and translations, rotations, cutout ^47^ as well as Gaussian blurring. Such augmented images were passed forward to the segmenter during training, along with equivalently transformed ground-truth instance segmentation masks; these segmentation masks were generated by manual data annotation followed by boot-strapping and manual curation. The internal parameters of Mask-RCNN were optimised to match the task at hand, consisting of modifications to its Region Proposal Network (RPN) and Non-Maximum Suppression parameters. The segmenter training was then optimised via a grid-search, keeping the model that performed best on validation data.

In the end, the best performing model was trained using an initial learning rate of 0.003 and batch size of 2 with the Adam ^48^ optimizer, using a momentum of 0.9 and weight decay of 0.001. The final model was trained in 4 consecutive steps, starting from initial weights trained on the MS COCO dataset ^49^. In the first step, the model ‘top’, consisting of the RPN and a second stage classifier and mask regressor were trained for 50 epochs at the initial learning rate, with other weights frozen. In the second step, the entire network was trained together, including the feature-encoding backbone, at the initial learning rate for another 50 epochs. In the 3rd step, the ‘top’ was fine-tuned for another 50 epochs, this time using 10 % of the initial learning rate (0.0003). Finally, the entire network was fine-tuned at 10 % of the initial learning rate for the final 50 epochs.

The Mask-RCNN model was adapted from a standard implementation ^50^.

### Deep Learning: classification

To generate training data for the DenseNet121 ^39^ classifier, both channels of every FoV acquired were used. The 30 frames of each FoV were averaged separately for both channels and used to construct RGB images, with Nile Red signal in the red channel and DAPI signal in the green. The DAPI channel was registered automatically to the Nile Red channel using cross-correlation ^51^ to correct for any drift between the channels. Individual cells were extracted from assembled images using the ground-truth instance segmentation masks that were used to train the segmenter. All cells were then resized to a common size of 64 by 64 pixels by zero-padding in either dimension if below the target size, or resized down to target size if above. To compensate for differences in staining and illumination, histogram equalization was applied to every cell, independently for each channel, within the segmentation mask only. Cells were then augmented on-the-fly using a random sequence of affine transformations, followed by a random sequence of intensity augmentations to increase robustness against experimental variation – these include a unsharp masking, random brightness modifier in HSB colour space, addition of Gaussian-distributed noise, channel misalignment and random Gaussian blurring. The classifier training was then optimized via grid-search, keeping the model that achieved the best accuracy on validation data.

To train the binary ciprofloxacin resistant/sensitive, individual untreated and ciprofloxacin treated MG1655 sensitive cells only were used; analogously for the other antibiotics.

The classifier was implemented in Keras^52^ version 2.2.4.

### Deep Learning: saliency mapping

To produce attention heatmaps over example classification inputs, we calculated the gradient of the output category with respect to the input single-cell image. We propagated positive gradients for positive activations only ^40^, and visualized the absolute value of the gradient.

### Segmentation metrics

The quality of Mask-RCNN segmentation was analysed using Precision-Recall curves using the bounding boxes of detections and ground truth segmentations to compare performance at various IoU thresholds.

### Classification metrics

Classification metrics of binary resistant-susceptible classifiers are presented as a confusion matrix, which displays the True Positive (TP), True Negative (TN), False Positive (FP) and False Negative (FN) counts in each class. From these, per class sensitivity, specificity and accuracy can be calculated as follows:

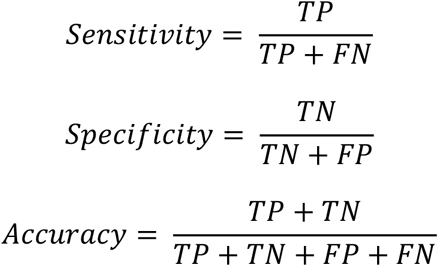

### Dose-Response Model fitting

To model the ratio of cells classified as susceptible as a function of treatment concentration, we use non-linear least squares to fit a generalized logistic function of the following form:

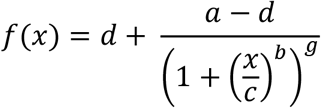

where *a* and *d* are the lower and upper asymptotes, *b* is the scale parameter, *c* is the x-coordinate of the inflexion point, and *g* is the asymmetry parameter. The confidence bands (CB) of the fit can be calculated directly from the covariance matrix of the fit:

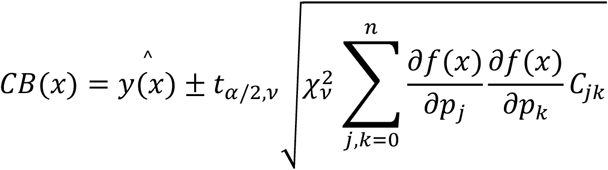

where 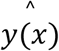is the best-fit estimate at *x, t*_*α*/2,*v*_ is the upper α/2 critical value for the t-distribution with N-n degrees of freedom, ν is the degrees of freedom, 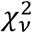is the reduced chi-square of the fit, *C* is the covariance matrix, *p* are the best-fit parameters and *f(x)* is the generalized logistic function.

### Statistics

The ratios of susceptible cells found and classified in untreated and ciprofloxacin treated clinical isolates were analysed using Tukey’s range test, computed pairwise for untreated and treated samples of each clinical isolate, and separately for each isolate. Three biological repeats were analysed for each isolate, producing a set of three susceptible cell ratios (n=3) per isolate, per biological repeat. The total number of cells that contributed to each ratio is displayed in Fig. S14. The significance level of the test was set at 0.05.

### Ethics

Ethical approval for the use of clinical samples and isolates processed by the John Radcliffe Hospital microbiology laboratory in the development of diagnostic assays was granted by the UK’s Health Research Authority (London - Queen Square Research Ethics Committee [REC reference: 17/LO/1420]).

## Data Availability

The data that support the findings of this study, including micrographs of treated and untreated cells, ground-truth annotations and trained models used to evaluate results are available from the corresponding author upon reasonable request.

## ACKNOWLEDGEMENTS

We thank Mantas Krisciunas whose early exploratory work provided motivation for this project. This work was supported by an Oxford Martin School (by the establishment of the Oxford Martin School Programme on Antimicrobial Resistance Testing; to ANK, NS, CN, DC, and MA), by Wellcome Trust grant 110164/Z/15/Z (to ANK) and by UK Biotechnology and Biological Sciences Research Council grants BB/N018656/1 and BB/S008896/1 (to ANK). The work was also supported by an UK Engineering and Physical Sciences Research Council funded scholarship (to AZ), part of the Oxford-Nottingham Biomedical Imaging Centre for Doctoral Training grant EP/L016052/1. The research was additionally supported by the National Institute for Health Research (NIHR) Health Protection Research Unit in Healthcare Associated Infections and Antimicrobial Resistance (NIHR200915) at the University of Oxford in partnership with United Kingdom Health Security Agency (UKHSA) and by the NIHR Oxford Biomedical Research Centre. NS is an NIHR Oxford BRC Senior Research Fellow. The views expressed in this publication are those of the authors and not necessarily those of the NHS, NIHR, the Department of Health or Public Health England. Figures 1 and S5 accompanying this manuscript were created with Biorender.com.

## Author contribution statement

ANK conceived and supervised the project. AZ, HES, and ANK designed experiments. MS designed experiments and collected preliminary data at early stages of the project. AZ carried out experiments, developed, trained and optimized computational models, analyzed data. PT assisted with data analysis, CF and HES assisted with experiments. LB and SO provided clinical isolates used in this work. CN, NS, MA, HES, MS, DC and ANK contributed to the interpretation of the results and shaped the strategic direction of the project. AZ and ANK wrote the manuscript. All authors discussed the results and contributed to the final manuscript.

## Code availability statement

All code used to generate the results is available publicly on the Kapanidis Laboratory GitHub account, accessible at: *https://github.com/KapanidisLab/Deep-Learning-and-Single-Cell-Phenotyping-for-Rapid-Antimicrobial-Susceptibility-Testing*. Detailed guidance on using the code is available upon reasonable request from AZ.

## REFERENCES

1. Murray, C. J. et al. Global burden of bacterial antimicrobial resistance in 2019: a systematic analysis. Lancet 399, 629–655 (2022).

2. Munita, J. M. & Arias, C. A. Mechanisms of Antibiotic Resistance. Microbiol Spectr. 4, 1–37 (2016).

3. Wouters, O. J., McKee, M. & Luyten, J. Estimated Research and Development Investment Needed to Bring a New Medicine to Market, 2009-2018. JAMA - Journal of the American Medical Association vol. 323 844–853 at https://doi.org/10.1001/jama.2020.1166 (2020).

4. Jackson, N., Czaplewski, L. & Piddock, L. J. V. Discovery and development of new antibacterial drugs: Learning from experience? Journal of Antimicrobial Chemotherapy vol. 73 1452–1459 at https://doi.org/10.1093/jac/dky019 (2018).

5. Schweitzer, V. A. et al. The quality of studies evaluating antimicrobial stewardship interventions: a systematic review. Clinical Microbiology and Infection vol. 25 555–561 at https://doi.org/10.1016/j.cmi.2018.11.002 (2019).

6. Jorgensen, J. H. & Ferraro, M. J. Antimicrobial susceptibility testing: a review of general principles and contemporary practices. Clin. Infect. Dis. 49, 1749–55 (2009).

7. Khan, Z. A., Siddiqui, M. F. & Park, S. Current and Emerging Methods of Antibiotic Susceptibility Testing. Diagnostics 9, (2019).

8. Vasala, A., Hytönen, V. P. & Laitinen, O. H. Modern Tools for Rapid Diagnostics of Antimicrobial Resistance. Frontiers in Cellular and Infection Microbiology vol. 10 at https://doi.org/10.3389/fcimb.2020.00308 (2020).

9. Khan, Z. A., Siddiqui, M. F. & Park, S. Current and emerging methods of antibiotic susceptibility testing. Diagnostics vol. 9 at https://doi.org/10.3390/diagnostics9020049 (2019).

10. Mulroney, K. et al. Same-day confirmation of infection and antimicrobial susceptibility profiling using flow cytometry. eBioMedicine 82, 104145 (2022).

11. Rousseau, A. N. et al. Fast Antibiotic Susceptibility Testing via Raman Microspectrometry on Single Bacteria: An MRSA Case Study. ACS Omega (2021) doi:10.1021/acsomega.1c00170.

12. Liu, M. et al. Single-Cell Identification, Drug Susceptibility Test, and Whole-genome Sequencing of Helicobacter pylori Directly from Gastric Biopsy by Clinical Antimicrobial Susceptibility Test Ramanometry. Clin. Chem. 68, 1064–1074 (2022).

13. Hsieh, K., Mach, K. E., Zhang, P., Liao, J. C. & Wang, T. H. Combating Antimicrobial Resistance via Single-Cell Diagnostic Technologies Powered by Droplet Microfluidics. Acc. Chem. Res. 55, 123–133 (2022).

14. Spencer, D. C. et al. A fast impedance-based antimicrobial susceptibility test. Nat. Commun. 11, (2020).

15. Li, H. et al. Adaptable microfluidic system for single-cell pathogen classification and antimicrobial susceptibility testing. Proc. Natl. Acad. Sci. U. S. A. 116, 10270–10279 (2019).

16. Baltekin, Ö., Boucharin, A., Tano, E., Andersson, D. I. & Elf, J. Antibiotic susceptibility testing in less than 30 min using direct single-cell imaging. Proc. Natl. Acad. Sci. 114, 9170–9175 (2017).

17. Roth, B. L., Poot, M., Yue, S. T. & Millard, P. J. Bacterial Viability and Antibiotic Susceptibility Testing with SYTOX Green Nucleic Acid Stain. Applied and Environmental Microbiology vol. 63 (1997).

18. Choi, J. et al. A rapid antimicrobial susceptibility test based on single-cell morphological analysis. Sci. Transl. Med. 6, (2014).

19. Ren, Y. et al. Prediction of antimicrobial resistance based on whole-genome sequencing and machine learning. Bioinformatics 38, 325–334 (2022).

20. Madrigal, P. et al. Machine learning algorithm to characterize antimicrobial resistance associated with the International Space Station surface microbiome. Microbiome 10, 1–12 (2022).

21. Ciloglu, F. U. et al. Drug-resistant Staphylococcus aureus bacteria detection by combining surface-enhanced Raman spectroscopy (SERS) and deep learning techniques. Sci. Rep. 11, 1–12 (2021).

22. Thrift, W. J. et al. Deep learning analysis of vibrational spectra of bacterial lysate for rapid antimicrobial susceptibility testing. ACS Nano 14, 15336–15348 (2020).

23. Al-Shaebi, Z., Uysal Ciloglu, F., Nasser, M. & Aydin, O. Highly Accurate Identification of Bacteria’s Antibiotic Resistance Based on Raman Spectroscopy and U-Net Deep Learning Algorithms. ACS Omega 7, 29443–29451 (2022).

24. Weis, C. V., Jutzeler, C. R. & Borgwardt, K. Machine learning for microbial identification and antimicrobial susceptibility testing on MALDI-TOF mass spectra: a systematic review. Clin. Microbiol. Infect. 26, 1310–1317 (2020).

25. Kandavalli, V., Karempudi, P., Larsson, J. & Elf, J. Rapid antibiotic susceptibility testing and species identification for mixed samples. Nat. Commun. 13, 1–8 (2022).

26. Charnot-Katsikas, A. et al. Use of the Accelerate Pheno System for Identification and Antimicrobial Susceptibility Testing of Pathogens in Positive Blood Cultures and Impact on Time to Results and Workflow. J. Clin. Microbiol. 56, (2017).

27. Iriya, R. et al. Rapid Antibiotic Susceptibility Testing Based on Bacterial Motion Patterns with Long Short-Term Memory Neural Networks. IEEE Sens. J. 20, 4940–4950 (2020).

28. Yu, H. et al. Phenotypic Antimicrobial Susceptibility Testing with Deep Learning Video Microscopy. Anal. Chem. 90, 6314–6322 (2018).

29. Bakshi, S. et al. Nonperturbative Imaging of Nucleoid Morphology in Live Bacterial Cells during an Antimicrobial Peptide Attack. Appl. Environ. Microbiol. 80, 4977–4986 (2014).

30. Spahn, C. et al. DeepBacs for multi-task bacterial image analysis using open-source deep learning approaches. Commun. Biol. 5, (2022).

31. Nonejuie, P., Burkart, M., Pogliano, K. & Pogliano, J. Bacterial cytological profiling rapidly identifies the cellular pathways targeted by antibacterial molecules. Proc. Natl. Acad. Sci. U. S. A. 110, 16169–16174 (2013).

32. Quach, D. T., Sakoulas, G., Nizet, V., Pogliano, J. & Pogliano, K. Bacterial Cytological Profiling (BCP) as a Rapid and Accurate Antimicrobial Susceptibility Testing Method for Staphylococcus aureus. EBioMedicine 4, 95–103 (2016).

33. Sridhar, S. et al. High-Content Imaging to Phenotype Antimicrobial Effects on Individual Bacteria at Scale. mSystems 6, (2021).

34. Araújo-Bazán, L., Ruiz-Avila, L. B., Andreu, D., Huecas, S. & Andreu, J. M. Cytological profile of antibacterial FtsZ inhibitors and synthetic peptide MciZ. Front. Microbiol. 7, (2016).

35. Htoo, H. H. et al. Bacterial Cytological Profiling as a Tool To Study Mechanisms of Action of Antibiotics That Are Active against Acinetobacter baumannii. Antimicrob. Agents Chemother. 63, (2019).

36. Lioy, V. S. et al. Multiscale Structuring of the E. coli Chromosome by Nucleoid-Associated and Condensin Proteins. Cell 172, 771-783.e18 (2018).

37. Vickridge, E., Planchenault, C., Cockram, C., Junceda, I. G. & Espéli, O. Management of E. coli sister chromatid cohesion in response to genotoxic stress. Nat. Commun. 8, (2017).

38. He, K., Gkioxari, G., Dollar, P. & Girshick, R. Mask R-CNN. in International Conference on Computer Vision 2961–2969 (2017).

39. Huang, G., Liu, Z., Van Der Maaten, L. & Weinberger, K. Q. Densely Connected Convolutional Networks. Proc. IEEE Conf. Comput. Vis. Pattern Recognit. 4700–4708 (2017).

40. Springenberg, J. T., Dosovitskiy, A., Brox, T. & Riedmiller, M. Striving for simplicity: The all convolutional net. Proc. IEEE Conf. Comput. Vis. Pattern Recognit. 1–14 (2015).

41. EUCAST. The European Committee on Antimicrobial Susceptibility Testing. Breakpoint tables for interpretation of MICs and zone diameters. Version 12.0, 2022. http://www.eucast.org.

42. Yoon, C. H. et al. Mortality risks associated with empirical antibiotic activity in Escherichia coli bacteraemia: an analysis of electronic health records. J. Antimicrob. Chemother. 77, 2536–2545 (2022).

43. Opota, O., Croxatto, A., Prod’hom, G. & Greub, G. Blood culture-based diagnosis of bacteraemia: State of the art. Clin. Microbiol. Infect. 21, 313–322 (2015).

44. Lipworth, S. et al. Ten-year longitudinal molecular epidemiology study of Escherichia coli and Klebsiella species bloodstream infections in Oxfordshire, UK. Genome Med. 13, 1–13 (2021).

45. Bortolaia, V. et al. ResFinder 4.0 for predictions of phenotypes from genotypes. J. Antimicrob. Chemother. 75, 3491–3500 (2020).

46. Seemann, T. Abricate. at https://github.com/tseemann/abricate (2020).

47. DeVries, T. & Taylor, G. W. Improved Regularization of Convolutional Neural Networks with Cutout. ArXiv (2017).

48. Kingma, D. P. & Ba, J. L. Adam: A method for stochastic optimization. 3rd Int. Conf. Learn. Represent. ICLR 2015 - Conf. Track Proc. 1–15 (2015).

49. Lin, T. Y. et al. Microsoft COCO: Common objects in context. Lect. Notes Comput. Sci. (including Subser. Lect. Notes Artif. Intell. Lect. Notes Bioinformatics) 8693 LNCS, 740–755 (2014).

50. Abdulla, W. Mask R-CNN for object detection and instance segmentation on Keras and TensorFlow. at https://github.com/matterport/Mask_RCNN (2017).

51. Maes, F., Loeckx, D., Vandermeulen, D. & Suetens, P. Image registration using mutual information. in Handbook of Biomedical Imaging: Methodologies and Clinical Research (eds. Paragios, N., Duncan, J. & Ayache, N.) 295–308 (Springer US, 2015). doi:10.1007/978-0-387-09749-7_16.

52. François, C. Keras. at https://keras.io (2015).

