## Supplemental Figures for "Deep Learning and Single Cell Phenotyping for Rapid Antimicrobial Susceptibility Testing"

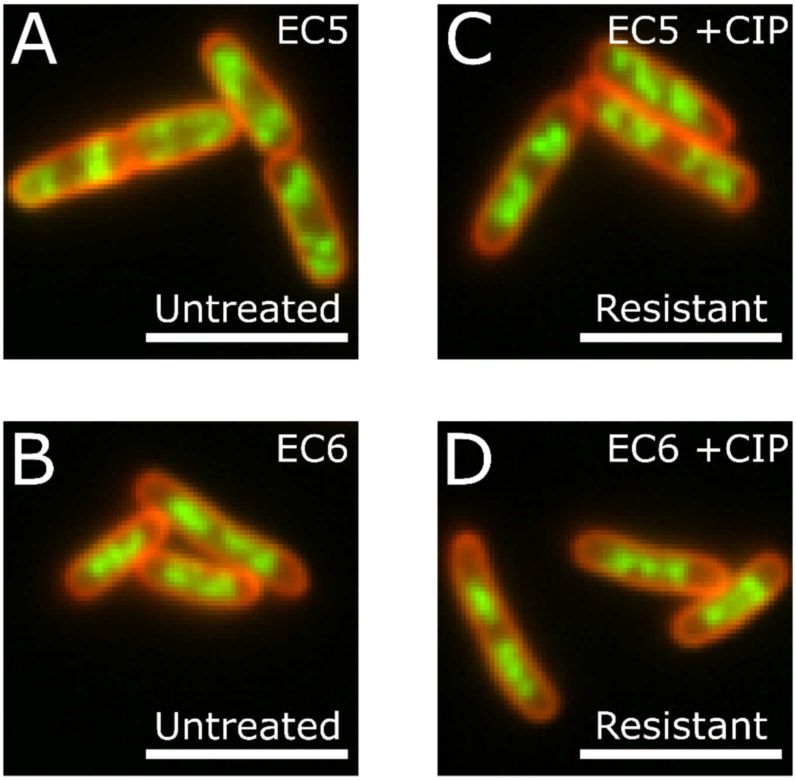


**Fig S1. Comparison of the resistant and untreated phenotype in ciprofloxacin resistant clinical isolates.** (A-B) Untreated phenotype in ciprofloxacin resistant EC5 and EC6. (C-D) Resistant phenotype in EC5 and EC6, after treatment with ciprofloxacin.

| Antibiotic | Code | EUCAST breakpoint (mg/L) | Treatment concentration (mg/L) | Treatment duration (min) |
|---|---|---|---|---|
| Ciprofloxacin | CIP | 0.5 | 10 | 30 |
| Gentamicin | GENT | 2 | 40 | 30 |
| Rifampicin | RIF | - | 100 | 30 |
| Co-amoxiclav | COAMOX | 8 | 160 | 60 |

**Fig S2. Treatment conditions and abbreviations used to generate susceptible phenotype training data in MG1655 *E. coli*.** Antibiotic column contains the full, standard name of the antibiotic. The code refers to the abbreviated names for these antibiotics used in this publication. The EUCAST breakpoints are the standardised thresholds used clinically for classifying specimens as either resistant or susceptible, via growth inhibition on agar plate, in our setting. No breakpoint has been established for the use of rifampicin in *E. coli*. The treatment concentration and duration are the antibiotic exposure parameters used to generate susceptible phenotypes in this study.

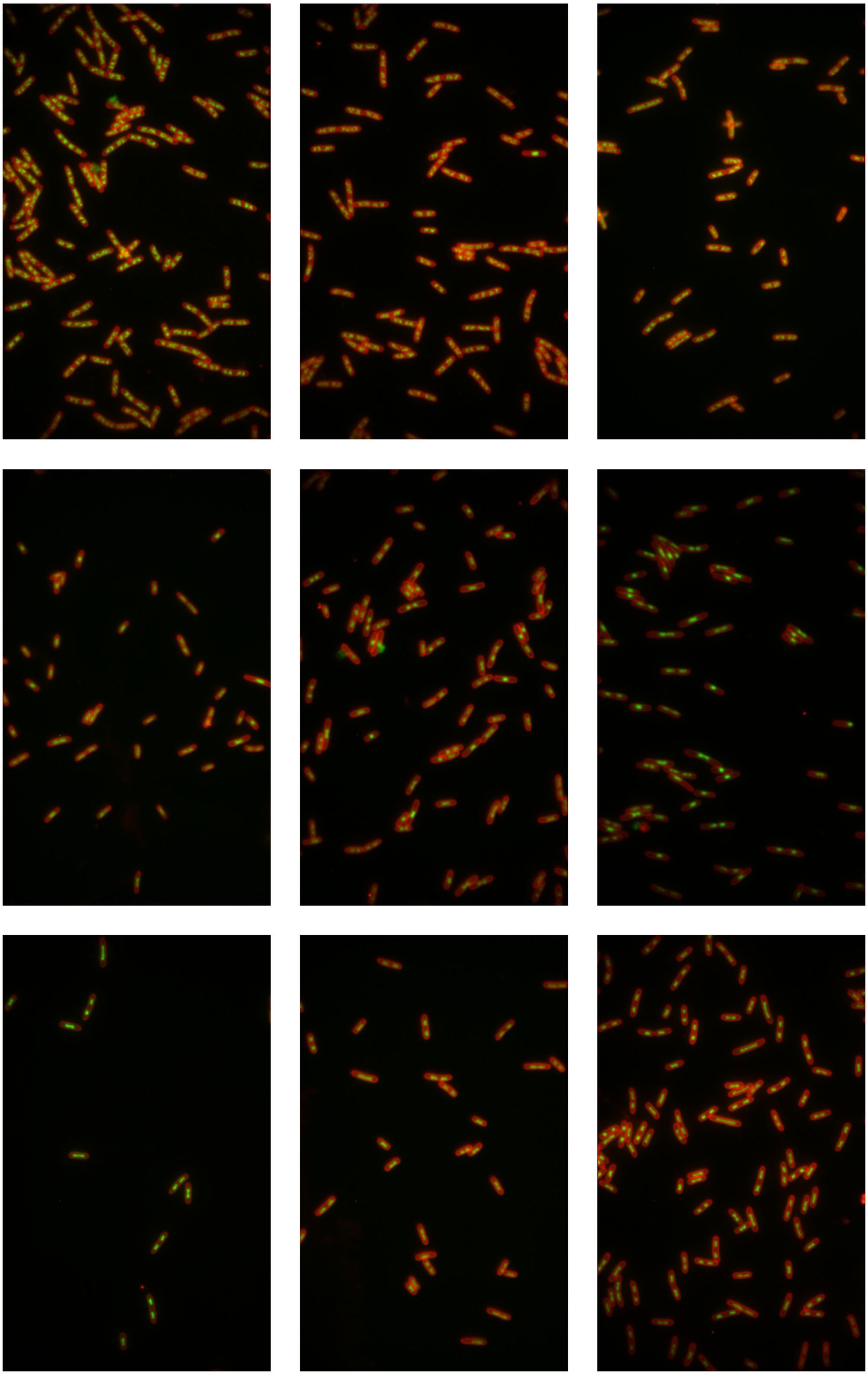

**Fig S3. Representative fields of view containing MG1655 *E. coli* cells showing the untreated, ciprofloxacin and gentamicin susceptible phenotypes used for classifier training.** (top row) 3 example fields of view showing the untreated phenotype, where no antibiotic was used. This phenotype was used to train the resistant class. (middle row) 3 example fields of view showing the ciprofloxacin susceptible phenotype, as a result of incubation with ciprofloxacin at concentration and duration shown in Fig S2. (bottom row) As above, but showing the gentamicin susceptible phenotype, the result of incubation with gentamicin.

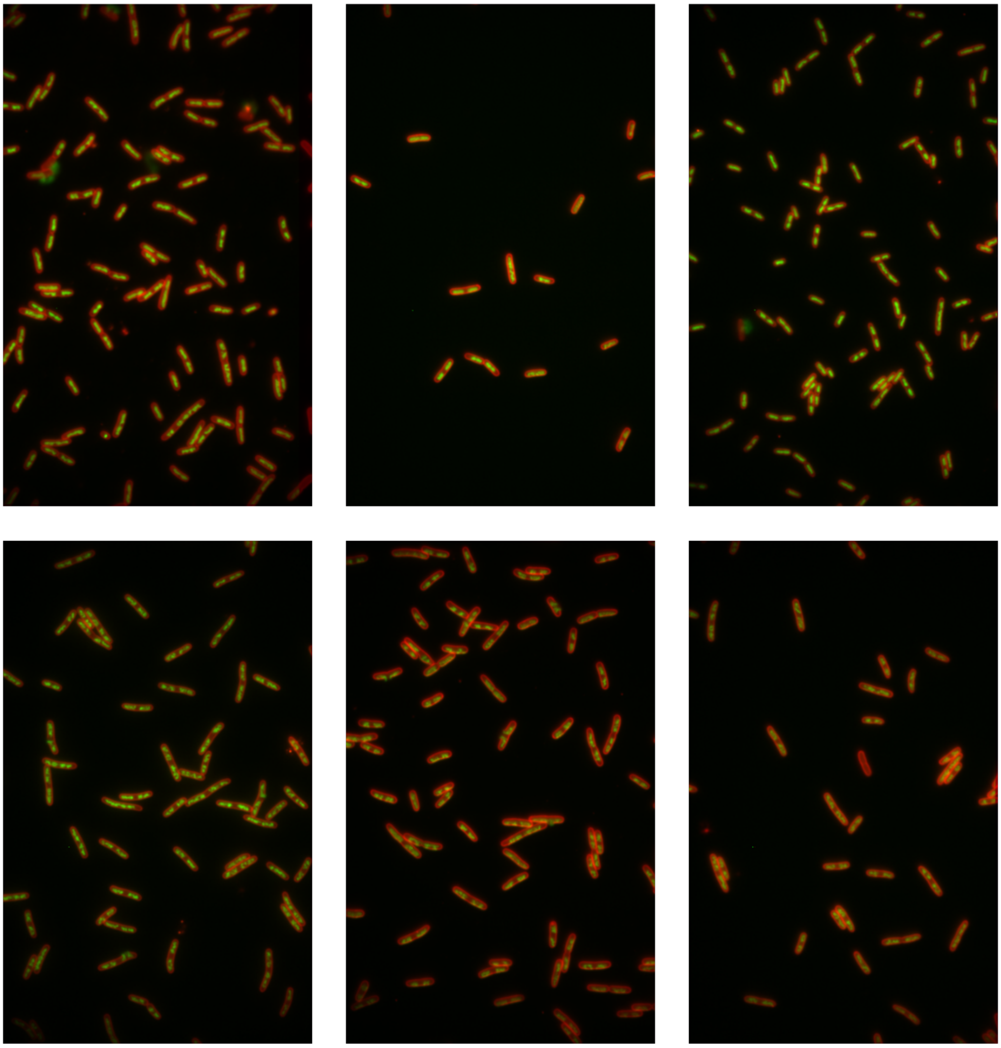

**Fig S4. Representative fields of view containing MG1655 *E.coli* cells showing the rifampicin and co-amoxiclav susceptible phenotypes used for classifier training.** (top row) 3 example fields of view showing the rifampicin susceptible phenotype, as a result of incubation with rifampicin at concentration and duration shown in Fig S2. (bottom row) As above, but showing the co-amoxiclav susceptible phenotype, the result of incubation with co-amoxiclav.

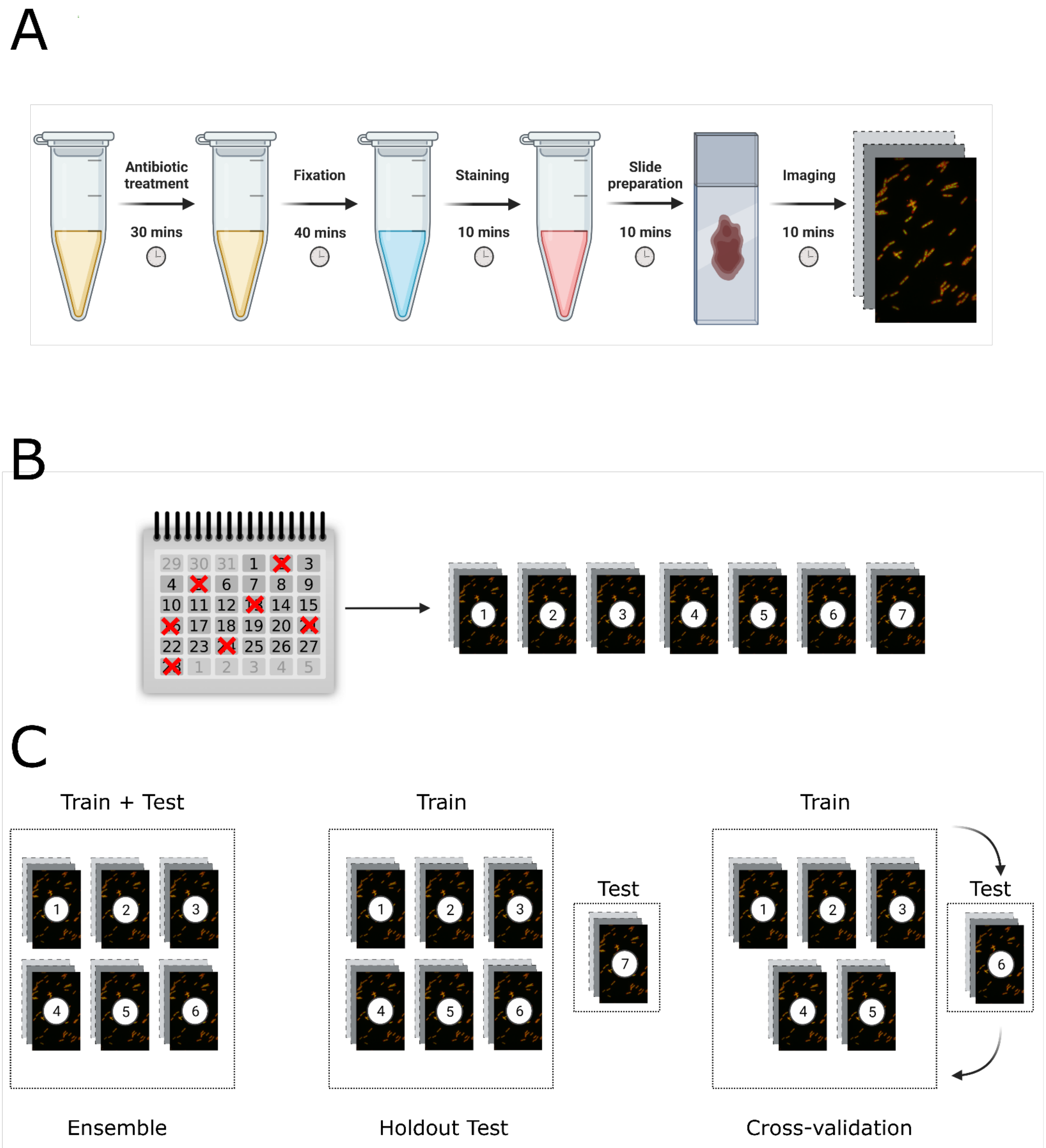


**Fig S5. Experimental and computational setup.** (A) Experimental setup. Live cells are treated with an antibiotic of choice, fixed with formaldehyde and permeabilised with ethanol. Fixed cells are stained with DAPI and Nile Red to stain the nucleoid and cell membrane respectively. The solution is placed on an agarose pad slide and imaged on an epifluorescence microscope. (B) 7 independent repeat experiments were conducted on 7 different days. 6 were used for training and cross-validation, one was the independent hold-out test. (C) Computational setup. (left) In the ensemble experiment, all non-edge cells from 6 different independent experiments are aggregated together, and divided into training and testing sets. (middle) In the holdout test experiment, the same number of cells per class per experiment was selected randomly from each of the 6 experiments, and used to train the model. The model was tested the same number of cells per class from a 7$^{th}$ repeat experiment – this experiment was not used for pipeline optimization or hyperparameter tuning. (right) In the cross-validation experiment, the same number of cells per class per experiment were randomly selected from each of the 6 experiments. One experiment was withheld to be used as the test set, and remaining 5 were used for training. The test experiment was rotated 6 times – the final result is the sum over 6 different models, each trained and tested on a different permutation of repeat experiments. Figure created with Biorender.com.

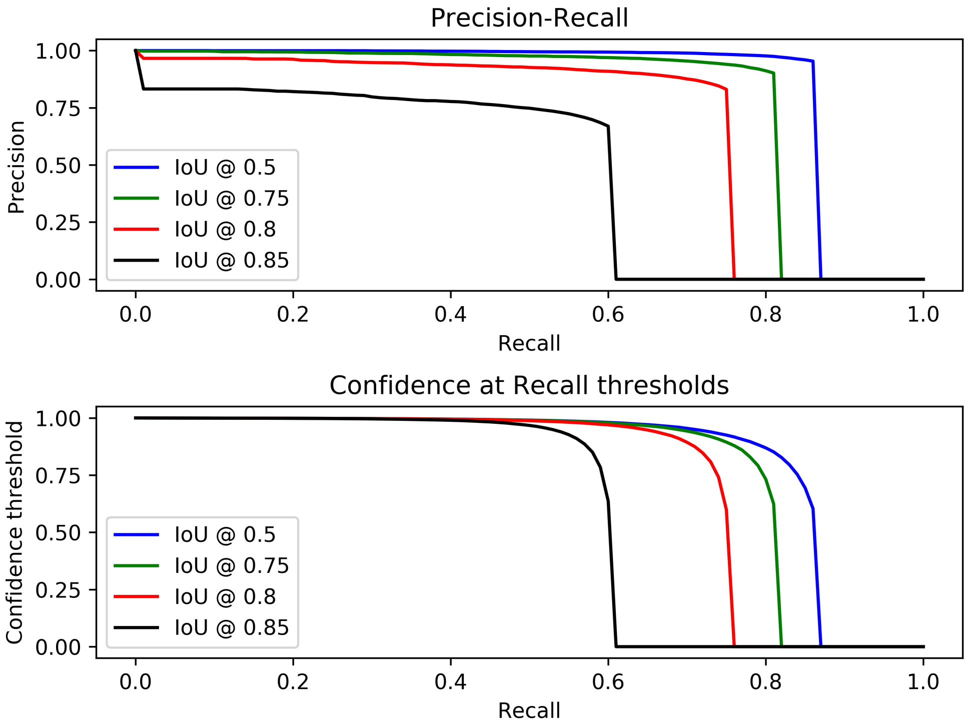


**Fig S6. Precision-Recall and corresponding confidence across a range of intersection over union (IoU) thresholds of the Mask-RCNN segmenter, evaluated on all micrographs from the holdout experiment.** Evaluated on 155 micrographs containing all of the phenotypes used in the study. (top) The precision-recall curve at 4 selected IoU thresholds. (bottom) The prediction confidence, corresponding to the precision-recall curve.

| ABX | TEST | TP | TN | FP | FN | Sensitivity | Specificity | Accuracy |
|---|---|---|---|---|---|---|---|---|
| CIP | ENSEMBLE | 2059 | 2653 | 197 | 143 | 94% | 93% | 93% |
| | CROSSVAL | 2739 | 2647 | 353 | 261 | 91% | 88% | 90% |
| | HOLDOUT | 422 | 467 | 33 | 78 | 84% | 93% | 89% |
| GENT | ENSEMBLE | 1629 | 714 | 160 | 57 | 97% | 82% | 92% |
| | CROSSVAL | 2627 | 2420 | 580 | 373 | 88% | 81% | 84% |
| | HOLDOUT | 468 | 459 | 41 | 32 | 94% | 92% | 93% |
| RIF | ENSEMBLE | 3808 | 2454 | 396 | 466 | 89% | 86% | 88% |
| | CROSSVAL | 2439 | 2482 | 518 | 561 | 81% | 83% | 82% |
| | HOLDOUT | 402 | 457 | 43 | 98 | 80% | 91% | 86% |
| COAMOX | ENSEMBLE | 957 | 622 | 252 | 210 | 82% | 71% | 77% |
| | CROSSVAL | 1586 | 1780 | 620 | 814 | 66% | 74% | 70% |
| | HOLDOUT | 352 | 322 | 78 | 48 | 88% | 80% | 84% |

**Fig S7. Full phenotype detection results in the *E. coli* MG1655 training strain across all antibiotics used in the study.** The ensemble, cross-validation (crossval) and holdout test (holdout) experiments were carried out on 4 different antibiotics (abx) used in the study. True positive (TP) is the number of treated cells correctly classified as susceptible. True negative (TN) is the number of untreated cells correctly classified in the resistant class (showing no response due to being untreated). False positive (FP) and false negative (FN) are the numbers of misclassified untreated cells and treated cells respectively. From these counts, the sensitivity, specificity and accuracy can be calculated.

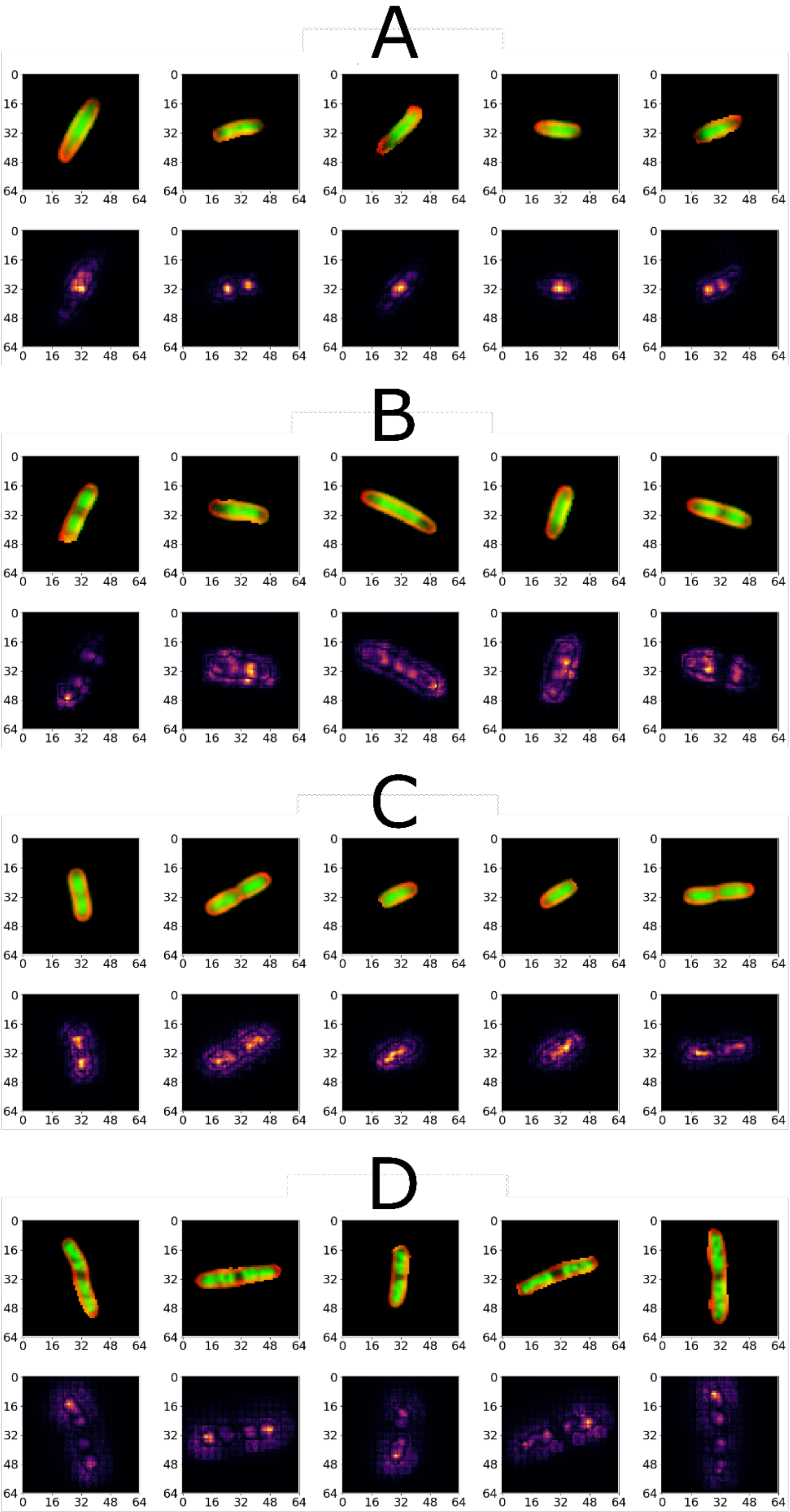


**Fig S8. Saliency mapping of randomly selected, correctly classified susceptible cells.** (A) (top) Randomly selected ciprofloxacin susceptible cells, after pre-processing steps, as presented to the classifier. All these cells were correctly classified as susceptible (bottom) Guided saliency maps corresponding to selected single-cell phenotypes, absolute value. Bright regions highlight areas that contributed most to the classification decision. (B) As above, but for gentamicin. (C) As above, but for rifampicin. (D) As above, but for co-amoxiclav.

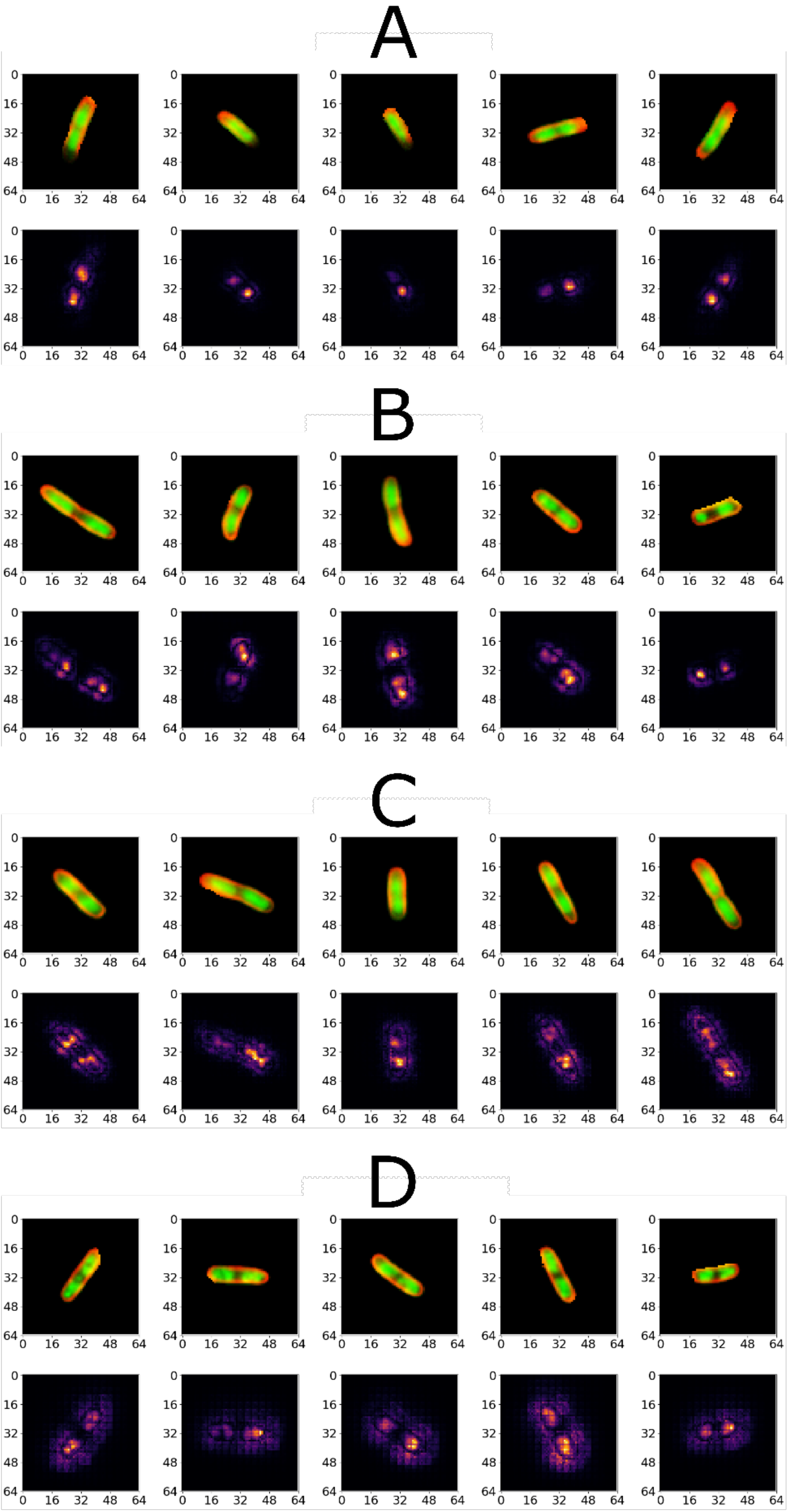


**Fig S9. Saliency mapping of randomly selected, misclassified susceptible cells.** (A) (top) Randomly selected, ciprofloxacin treated susceptible cells, after pre-processing steps, as presented to the classifier. All these cells were incorrectly classified as resistant. (bottom) Guided saliency maps corresponding to selected cells, absolute value. Bright spots highlight regions that contributed most to the classification decision. (B) As above, but for gentamicin. (C) As above, but for rifampicin. (D) As above, but for co-amoxiclav.

| Isolate name | Genotype | MIC (mg/L) |
|---|---|---|
| EC1 | *marR*$^{N3}$ | 0.008 |
| EC2 | *marR*$^{N3}$ | 0.03 |
| EC3 | *gyrA*$^{L83}$ *parC*$^{I80}$ | 0.5 |
| EC4 | *gyrA*$^{L83, N87}$ *parC*$^{I80}$ | 8 |
| EC5 | *gyrA*$^{L83, N87}$ *parC*$^{I80, V84}$ *parE*$^{L529}$ | 72 |
| EC6 | *gyrA*$^{L83, N87}$ *parC*$^{I80}$ *parE*$^{A458}$ | 108 |

**Fig S10. Table of clinical E. coli isolates used in the study.** Isolate name refers to the pseudorandom code used for isolates in the manuscript and figures. Genotype contains details of mutations associated with resistance to ciprofloxacin, detected by whole genome sequencing of the isolates, following the method described in Methods. The MIC column lists experimentally derived ciprofloxacin Minimum Inhibitory Concentrations (MIC), following the method described in Methods.

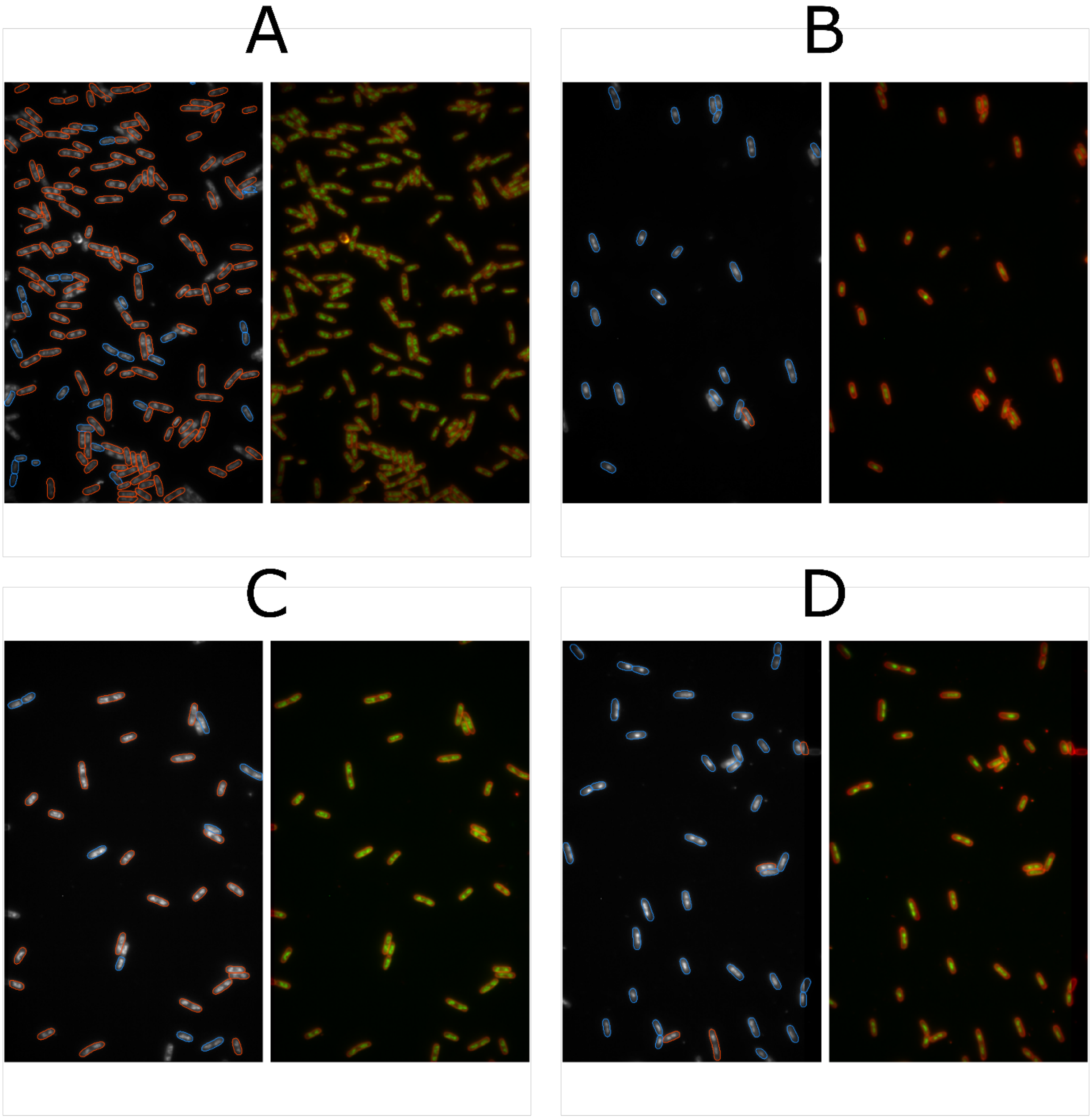


**Fig S11. Representative fields of view and detection overlays of ciprofloxacin-susceptible clinical isolates EC1 and EC2, in both untreated and ciprofloxacin treated conditions.** (A) (left) Detection overlay of a field of view of untreated EC2. Red detections are classified as resistant/untreated, blue detections are classified as susceptible. (right) Corresponding raw field of view. (B) As A, but for a ciprofloxacin treated EC2 field of view. (C) As A, but for untreated EC1. (D) As B, but for ciprofloxacin treated EC1.

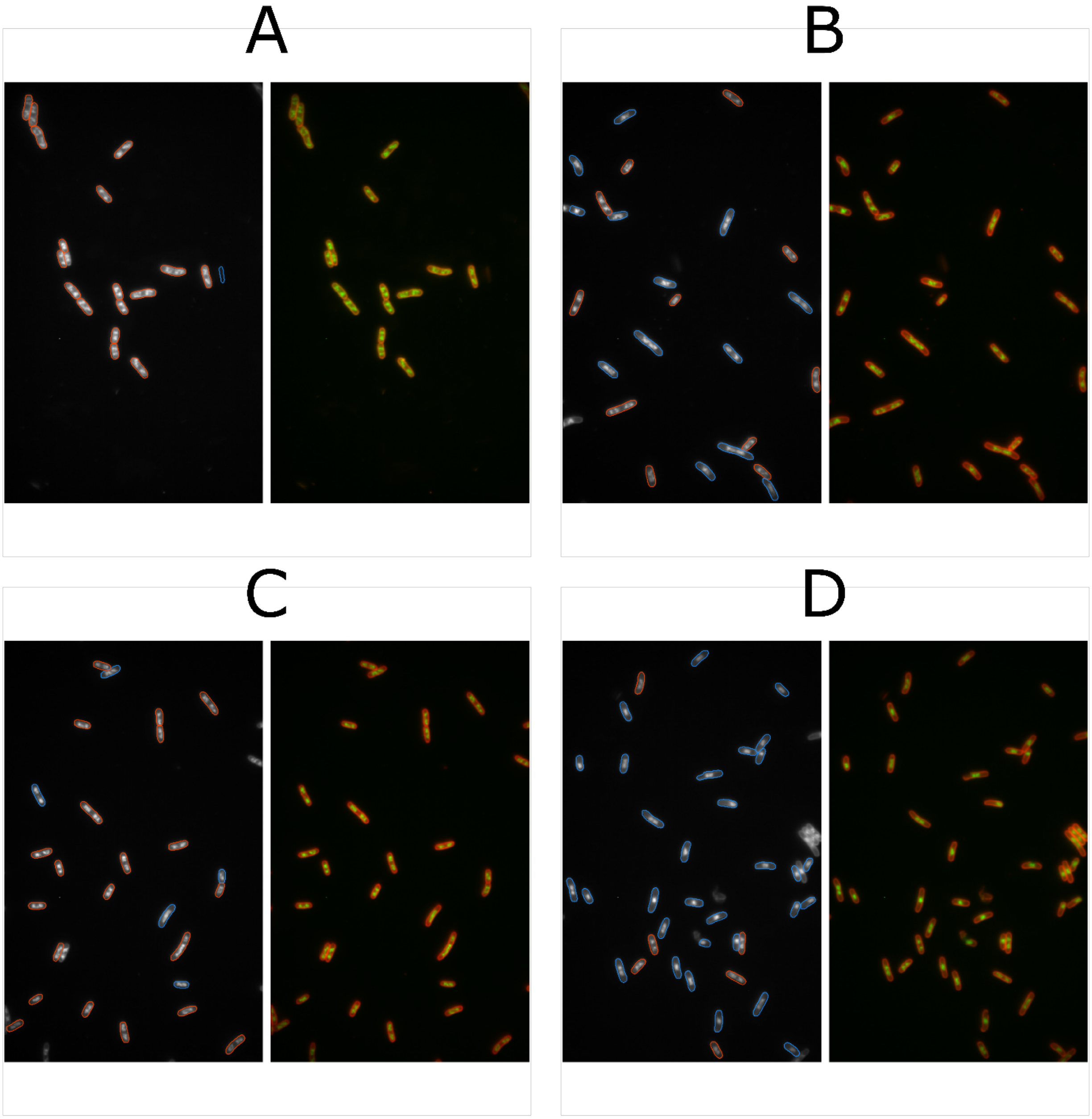


**Fig S12. Representative fields of view and detection overlays of ciprofloxacin-susceptible clinical isolates EC3 and EC4, in both untreated and ciprofloxacin treated conditions.** (A) (left) Detection overlay of a field of view of untreated EC4. Red detections are classified as resistant/untreated, blue detections are classified as susceptible. (right) Corresponding raw field of view. (B) As A, but for a ciprofloxacin treated EC4 field of view. (C) As A, but for untreated EC3. (D) As B, but for ciprofloxacin treated EC3.

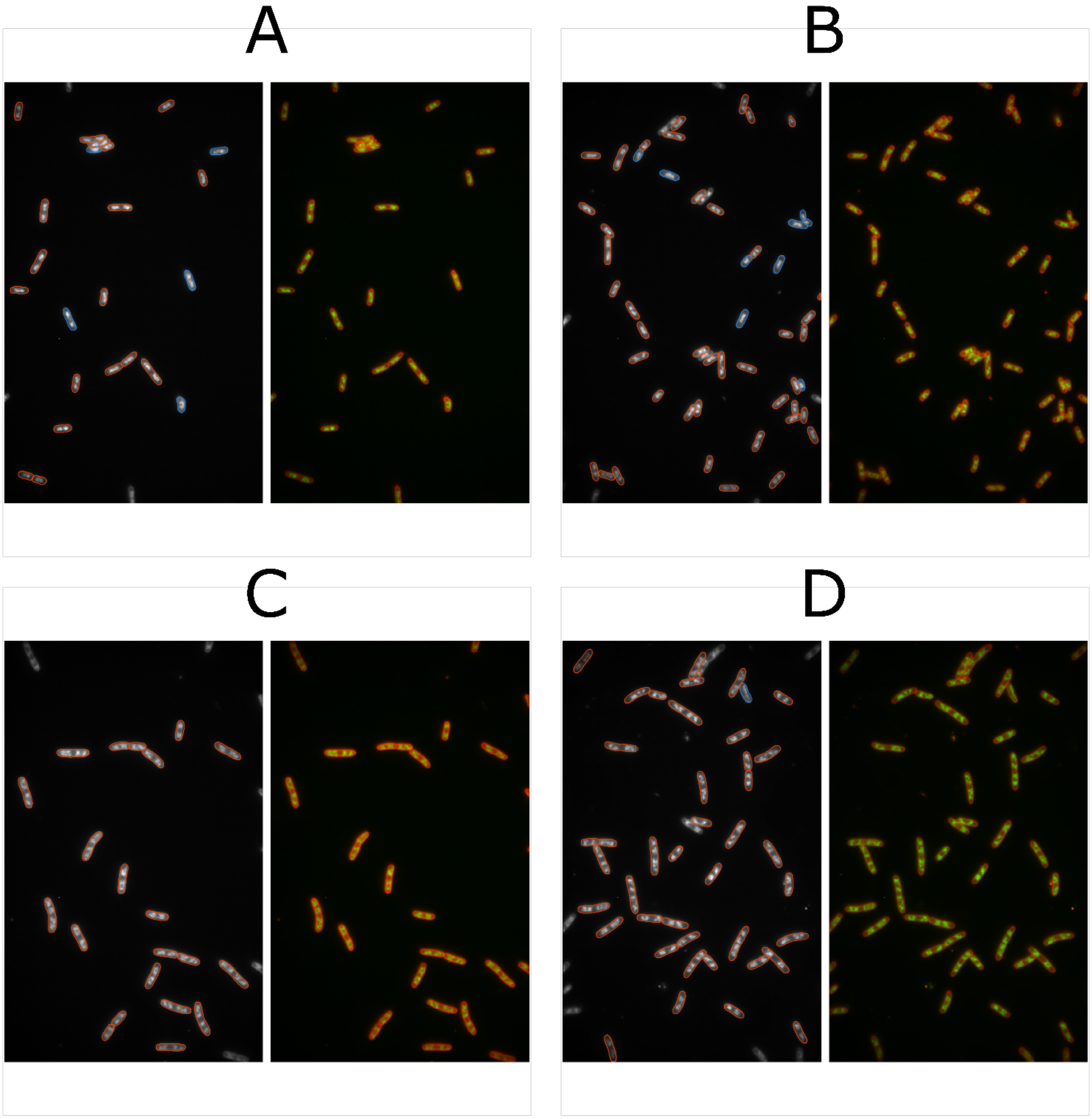


**Fig S13. Representative fields of view and detection overlays of ciprofloxacin-resistant clinical isolates EC5 and EC6, in both untreated and ciprofloxacin treated conditions.** (A) (left) Detection overlay of a field of view of untreated EC6. Red detections are classified as resistant, blue detections are classified as susceptible. (right) Corresponding raw field of view. (B) As A, but for a ciprofloxacin treated EC6 field of view. (C) As A, but for untreated EC5. (D) As B, but for ciprofloxacin treated EC5.

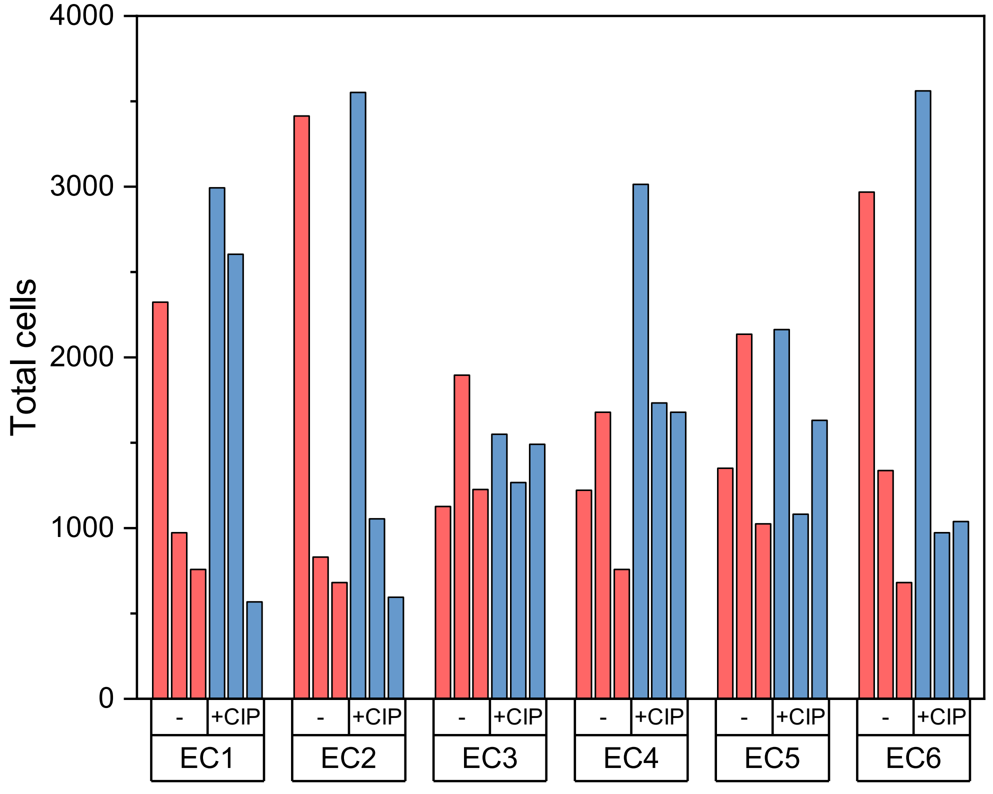


**Fig S14. Total numbers of cells detected in each of the biological replicate experiments used to generate Figure 5, in both untreated and ciprofloxacin treated conditions.** (top axis) Treatment condition. (bottom axis) Isolate code.

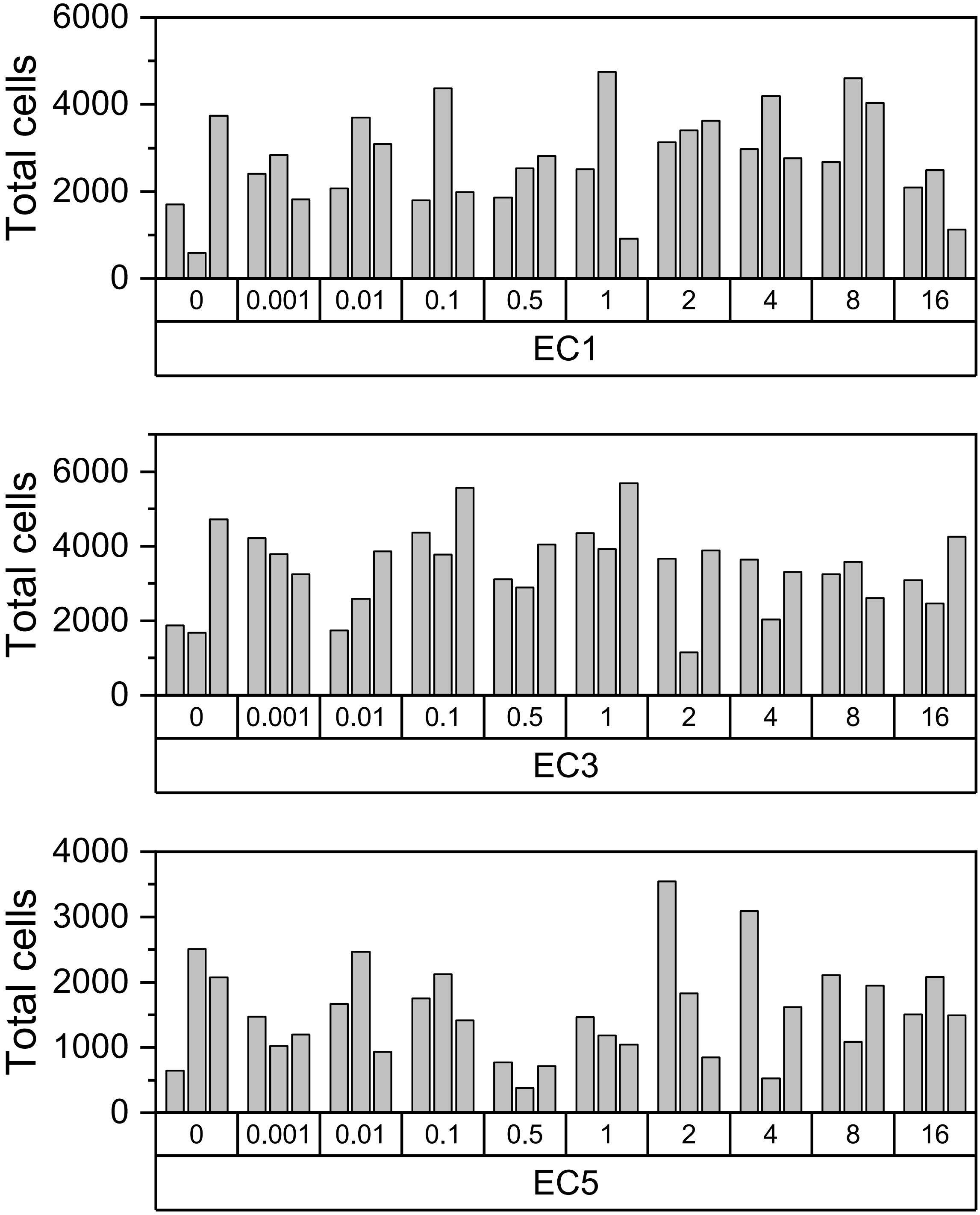


**Fig S15. Total numbers of cells detected in each of the biological replicate experiments used to generate Figure 6, in both untreated and ciprofloxacin treated conditions.** (top axis) Ciprofloxacin treatment concentration in mg/L. "0" indicates no antibiotic treatment. (bottom axis) Isolate code.
